# Dietary bioactives increase gut microbiome diversity and alter host and microbial metabolite profiles

**DOI:** 10.1101/2025.10.16.25338140

**Authors:** Anthony Duncan, Federico Bernuzzi, Jennifer Ahn-Jarvis, Liangzi Zhang, Daniela Segovia-Lizano, Maja Petrou, Janis Rebecca Bedarf, Duncan Ng, George M Savva, Jan Stanstrup, Lars Ove Dragsted, Falk Hildebrand, Maria H. Traka

## Abstract

**Objective:** Dietary bioactives have been associated with positive health effects such as cardiovascular health, or having anti-diabetic properties. Many bioactive compounds survive digestion arriving in the colon, where the gut microbiome can further metabolise them to forms more available to the host. Previous work has shown diets such as the Mediterranean diet, enriched in plant bioactives, affecting the composition and diversity of the gut microbiome. However, it is unclear which factors in such whole diet interventions underlie these changes, such as the bioactives themselves, the percentage of dietary fibres or macronutrient composition. In the Dietary BIoactives and Microbiome DivErsity (DIME) study we investigated the impact of a diet rich in a wide range of plant bioactives compared to a low bioactive diet with matched total fibre intake.

**Design:** We conducted a randomised crossover intervention trial to assess alterations to the gut microbiome, urinary and faecal metabolites in 20 healthy subjects receiving two-week high- or low-bioactive diets, separated by a 4-week washout period. Macronutrient and total fibre content was carefully matched between diets.

**Results:** Alpha diversity of both microbial taxa and function increased in the high bioactive diet. Microbiome compositions between participants appear more similar after the high bioactive diet than low, suggesting that the high bioactive diet selected for bacteria with a similar functional spectrum. Both faecal and urinary metabolites were strongly impacted by the diet intervention, including o-coumaric acid, theobromine, and secondary bile acids. Metabolite profiles were more strongly associated to the dietary intervention arms than microbiome profiles, suggesting that a diet high in bioactives may change the activity of the existing community despite only small shifts in community composition.

**Conclusion:** High intake of dietary bioactives leads to significant changes in both microbial and host metabolite profiles. However, the shift in microbiome composition was substantially less pronounced, highlighting the need of future studies to investigate metabolic activities instead of taxonomic compositions.

## Background

One of the key modulators of our gut microbial communities is the food we eat and studies have shown that diet interventions with a high proportion of plant products can rapidly change composition of gut microbiota as early as two days after a dietary change and increase gut microbial diversity[1–3]. However, food is a complex mixture of components, such as nutrients, complex carbohydrates including fibre, and dietary bioactive compounds, that all have the potential to affect gut microbiome communities in an additive, synergistic or antagonistic way, making it difficult to unravel the mechanistic relationship between food, gut microbiota and health.

Dietary bioactives are non-essential compounds in food which are abundant in nature and are responsible for the diverse colours and tastes of plant foods such as fruits and vegetables[8]. Some bioactives have been associated to health benefits, with evidence that dietary intakes of certain flavonoids can have beneficial effects on risk factors for cardiovascular disease such as LDL cholesterol and blood pressure[9,10]. The gut microbiome compositions can also be impacted by increased intake of individual dietary bioactives or sources of bioactives, amongst these, polyphenols, found in many red and black berries, carotenoids, found in orange fruits and vegetables, and sulfur-metabolites, rich in cruciferous and allium vegetables[11,12]. Many dietary bioactives are metabolised by the gut microbial community into forms that are more bioavailable both for the host and for the gut microbiome. In the case of polyphenols only ten per cent of polyphenols are absorbed within the upper gastrointestinal tract into the blood[13], while the remainder reach the colon. These are fermented via gut bacterial polyphenol metabolism into a range of secondary metabolites, such as phenolic acids, with some in vitro experiments of polyphenol digestion observing an increase in short chain fatty acid concentration[14].

Similarly, glucosinolates, present in cruciferous vegetables, rely predominantly on colon fermentation to produce bioactive compounds that have been shown to modulate gut microbiota[11,12]. Dietary bioactives may also modulate gut microbiome composition through antimicrobial properties, with evidence accumulating that bioactive compounds can inhibit the growth of pathogenic bacteria[15,16].

Previous studies designed to assess whether increasing our fruit and vegetable intake can positively affect our gut microbiota have focused on interventions using either a single food or food extract[17–20] often at doses not representative of dietary intakes. Others have undertaken a whole diet intervention, e.g. Mediterranean diet[21–23], or vegan diet[3], which changes not only dietary bioactives but also macro- and micronutrient composition, that in themselves can profoundly influence the gut microbiome[24–26]. Plants are sources of naturally occurring dietary fibre, and consequently plant rich dietary interventions often represent an increase in fibre intake over habitual diets. Dietary fibre intake can significantly alter the abundance of taxa such as Bifidobacteria in faecal microbiome[27], as well being associated with increased faecal butyrate concentrations[28]. Further, variation in the type of fibre can select for differing species in in vitro experiments[29]. In whole diet interventions. In whole diet interventions, the effects of increasing plant micronutrient intake are challenging to distinguish from those driven by higher levels of dietary fibre where interventions increase both simultaneously.

In this study, we aimed to study the effects of dietary bioactives separately from those of fibre intake when assessing effect of whole diet on gut microbiome. We designed the Dietary BIoactives and Microbiome DivErsity (DIME) dietary intervention study to test the hypothesis that a diet delivering high levels of a diverse range of bioactives but a comparable amount of other nutrients, including fibre, would result in beneficial changes in gut microbiota composition and function.

## Results

### Participant Description

A total of 27 healthy participants (mean ± sem; 38.9 ± 2.4 y) gave their informed consent and 20 (8 men and 12 women) participants completed all study activities (Figure 1A). Three participants completed the first intervention before crossover but withdrew or were withdrawn before the second intervention (Supplementary Figure 1). Participants were normotensive and considered healthy based on anthropometric measurements (Supplementary Table 1) and screening glycated haemoglobin (HbA1c: 33.17 ± 0.74 mmol/mol) being within normal limits, and with no conditions or current medications which may affect their metabolism or digestion (see Methods).

**Figure 1:**
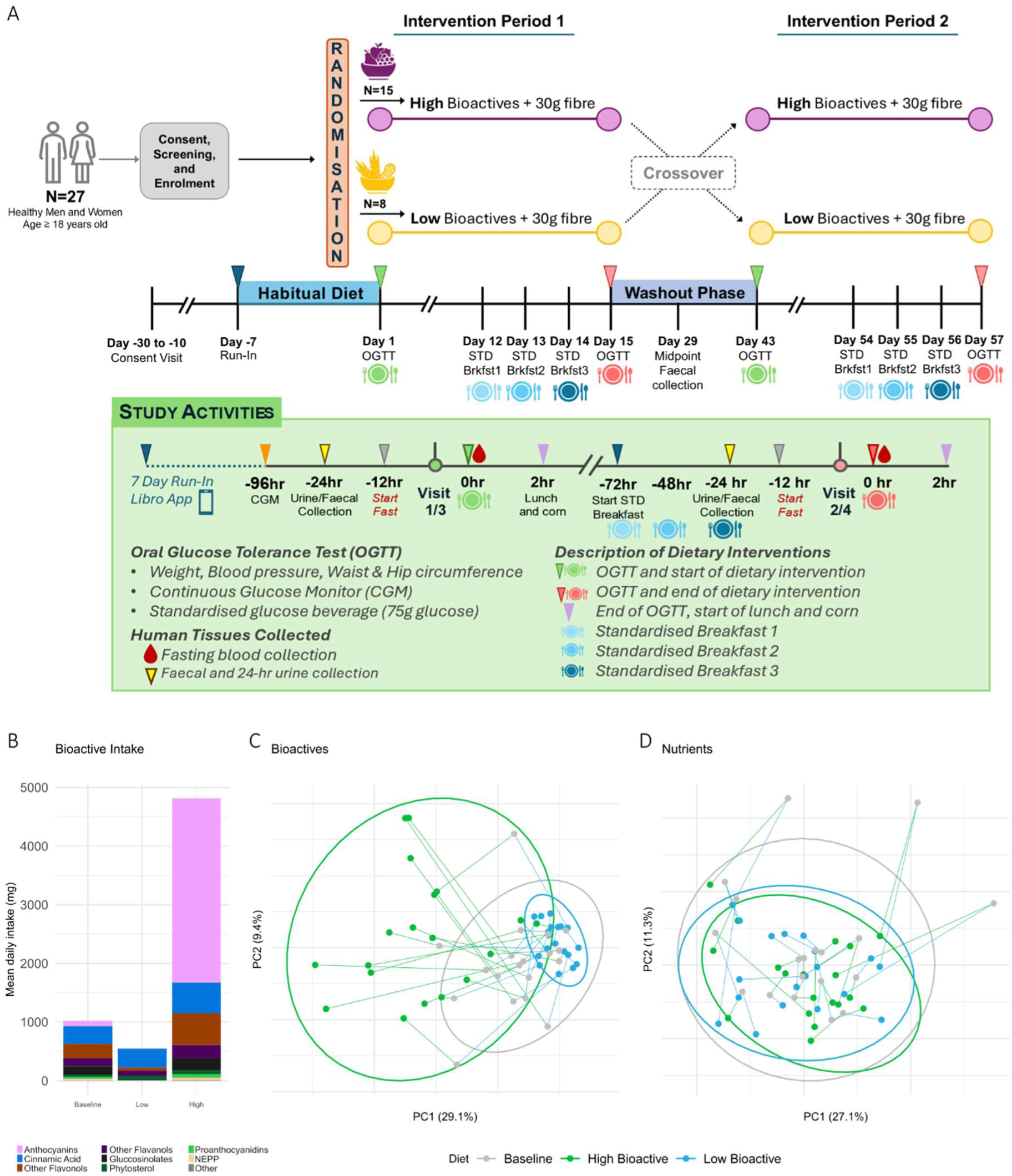
Dietary intake of bioactives and nutrients during the high and low bioactive diets, and baseline habitual diet. A) Diagram of study design. OGTT, Oral Glucose Tolerance Test; CGM, Continuous Glucose Monitor; STD, standard. B) Intake of bioactives by category in different interventions. Top 8 are shown, with the remaining summed under “Other”. Principal components analysis of C) bioactive intake and D) non-bioactive macro- and micronutrient intake during the two interventions and baseline. Lines connect samples relating to the same participant.

### Dietary Adherence and Reporting

Consistent with the instructions to participants, 91.5% (N=23) of them completed their 7-day diet entries during their habitual diet baseline (BLN) period. Likewise, participants completed their 14-day diet records during the low bioactive (LB) and high bioactive (HB) intervention periods (adherence as percentage of days recorded: LB 99.6% and HB 95.4%,). Since only 21% of the participants completed the 7-day diet record during the washout period, these diet records were omitted from the reporting. Additionally, records where the daily intake was less than 800 kcal or less than 50% of the participant’s average intake were removed from the analysis.

There were no significant differences in energy or total macronutrients intake between LB and HB; however, modest differences were observed between LB and HB in the various sugars (oligosaccharides, sucrose, glucose, and fructose) and fats (total, mono- and polyunsaturated). During the HB intervention there was an increase in total sugar intake, likely due to the increased intake of fruits, whereas during the LB diet there was a significant increase in total fat intake, likely due to the increased intake of mono- and polyunsaturated fats from nuts.

Consistent with the HB and LB diet instructions, differences between bioactives (anthocyanins, beta-carotene, ellagitannins/ellagic acid, glucosinolates, and lignans) were evident between LB and HB diets (Supplementary Table 2, Supplementary Figure 2).

Dietary intake of bioactives confirms diet adherence

The total daily bioactive intake was on average 1.08g during self-reported BLN diet, 4.97g during HB and 0.58g during LB diet interventions (Figure 1B). Anthocyanins were the most consumed bioactive class in the HB arm, with participants consuming 3.14g of anthocyanins compared to 0.007g in the LB arm and 0.094g at baseline (Supplementary Figure 2). Among other polyphenols, during the HB arm the participants increased their intake of flavanols, flavonols, cinnamic acid and derivatives, and ellagic acids, along with increased carotenoid and glucosinolate intakes. Higher intake of phytosterols, cysteine sulfoxides and capsaicinoids were observed in the LB arm (Supplementary Figure 2). Mean fibre intake in the baseline diets was 22.67g, increasing to 29.78g in the HB arm and 26.83g in the LB arm (Supplementary Figure 3).

To preclude confounding of the dietary analysis due to overall food intake, we first checked for correlation between mean daily energy intake and the summed mean daily intake of both the 27 bioactives and 107 nutrients quantified from food diaries. We found a mean positive correlation with total caloric intake for nutrients (𝑝 ≤ 0.001, Spearman’s 𝜌 = 0.36), but not for bioactives (𝑝 > 0.05, 𝜌 = −0.06). Next, we considered the relationship of mean daily caloric intake to mean daily intake of each bioactive and nutrient individually. Nutrient intake had a significantly stronger relationship to caloric intake than bioactives to caloric intake (comparing 𝑅^2^, 𝑝 < 0.001, Wilcoxon rank sum test, Supplementary Figure 4), suggesting that bioactive intake was less impacted by the amount of food consumed. To remove this potential confounder, bioactive and nutrient intakes were both adjusted in our subsequent analysis based on residuals of regression against total caloric intake[30].

Principal component analysis (PCoA) of dietary bioactives intake also confirmed that the three diets were significantly different (𝑝 ≤ 0.001, PERMANOVA Figure 1C), explaining 24% of variance, and post-hoc testing showed each combination of diets to be significantly different (𝑝 ≤ 0.001). In contrast, there was no grouping visible by intervention when looking at nutrient intake (Figure 2D). There was a difference in nutrient intake between diets (𝑝 ≤ 0.05, PERMANOVA) although explaining only 7% of variance, with differences between BLN and LB, and between LB and HB being significant (𝑝 ≤ 0.05) in post-hoc tests. Bioactive intake in the LB arm was less dispersed than either HB or baseline intakes (𝑝 < 0.01, betadisper, Fig. 2B), meaning that in terms of dietary bioactives the diets eaten during the LB arm were very similar for all subjects. In the HB arm bioactive intake was higher than BLN, but the dispersion between individuals was not different to BLN, showing that while bioactive intake increased there is no evidence that the bioactive composition of the HB diets became more dissimilar between individuals than it was at BLN.

**Figure 2:**
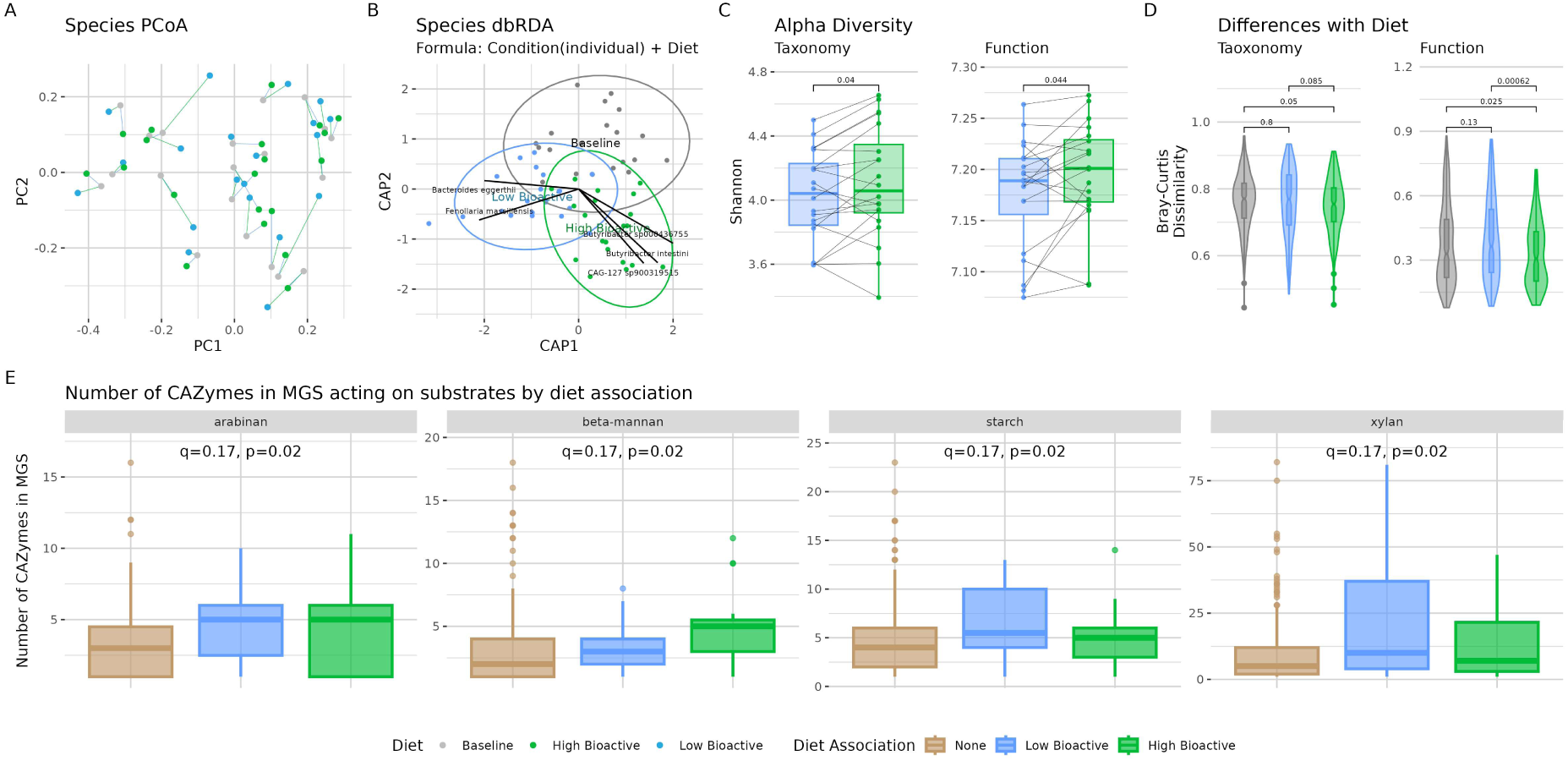
Taxonomic and functional composition of microbiome across dietary conditions. A) PCoA of species level relative abundance using Bray-Curtis dissimilarity. Samples from the same participant are joined using lines. B) Distance-based RDA of species level relative abundance using Bray-Curtis dissimilarity. The individual was used as conditioning term, and diet as a constraint. Species significantly correlated to the axes are indicated with lines. C) Alpha diversity measured using Shannon diversity index in HB and LB, p-values are from a Wilcoxon signed rank test. Taxonomy is at species level, and function using KO terms. D) Difference between samples in the post dietary intervention samples for HB and LB, and between all non-intervention samples for BLN (pre-diet and midpoint), with p-values from Wilcoxon rank sum test. Difference measured using Bray-Curtis dissimilarity on absolute abundance of species or KO terms. E) Counts of CAZymes utilising different substrates encoded by MGS associated with HB, LB, or not associated to diet in dbRDA. Results with Krusal-Wallis test p <= 0.05 displayed, though q >= 0.05 after BH adjustment for all.

### Consistent microbiome changes with high bioactive diet

From assembled metagenomes for all participants who completed the study (N=20), predicted genes were clustered at 95% identity to produce a gene catalogue with 4,164,116 representative genes. Binning metagenomic assemblies generated 1,848 MAGs (completeness ≥80%, contamination ≤5%), and clustering of the gene catalogue an additional 49 canopy clusters[31] passing the same completeness and contamination criteria. These MAGs and canopy clusters were dereplicated into 454 metagenomic species (MGS) that allow an unbiased, genome resolved view[32] on the taxonomic diversity in our metagenomes.

Interindividual differences dominated our dataset; accordingly, beta-diversity varied significantly between individuals (𝑝 < 0.001, PERMANOVA on Bray-Curtis dissimilarities, Figure 2A). Samples from the same individual clustered together in a principal coordinates analysis (Figure 2A) with no evident grouping by diet. Distance-based redundancy analysis (dbRDA) conditioned for participant and constrained on diet showed that 84% of the variance in dissimilarities could be accounted for by participant identity. On the dbRDA axes constrained on the two diet types (Figure 2B), HB and LB separated along dbRDA D1 (dimension 1), and HB + LB separated from BLN on D2.

Taxonomic alpha diversity showed a small increase after the HB-diet; Shannon index showed a statistically significant difference with a mean of 4.11 in HB, and 4.02 in LB (Figure 2C, p=0.04, Wilcoxon signed rank test), along with inverse Simpson (mean 34.1 HB; 28.4 LB, p=0.3) and richness (mean 187 HB; 182 LB, p=0.017, Supplementary Figure 5). Overall microbial load, estimated via flow cytometry (see Methods), did not vary significantly between the interventions or study visits (Supplementary Figure 6). We calculated pairwise beta diversity (Bray-Curtis dissimilarity) between all samples taken after the HB diet, to express how similar microbiomes from different individuals are to one another following a high bioactive diet; we did the same for LB, and non-intervention samples. Mean beta diversity between post-intervention samples within the HB arm was significantly lower that within non-intervention arms (mean HB 0.75; non-intervention 0.764, Figure 2D, p=0.05), suggesting that the microbiome composition became more similar during the HB intervention.

To understand compositional differences better, we calculated Enterosignature compositions of each sample, a model which describes the gut microbiome composition as a mixture of five bacterial guilds[33]. While the guilds in this model described our samples well (mean model fit 0.75), we found no significant changes between HB and LB. However, the Prevotella dominated Enterosignature ES_Prev decreased after the HB diet (𝑞 < 0.05, Wilcoxon signed rank test, Supplementary Figure 7), indicating that the genus Prevotella was likely negatively affected (or outcompeted) in HB conditions.

Using the dbRDA constrained on diet (Figure 2B), we identified MGS associated to each diet by setting thresholds on the angle between fitted MGS vectors and diet centroid and vector length (Methods, Supplementary Table 3), resulting in 39 HB and 39 LB associated MGS. *Bacteroides uniformis* and *Prevotella avicola* increased with the LB diet (determined by correlation to centroid vector), while the strongest association with the HB diet is *Phocaeicola massiliensis*, a species observed to encode enzymes involved in pectin and lignin degradation[34], fibers commonly found in fruits and plant cell walls respectively. Many of the other species whose vectors in the constrained axes are most similar to the HB centroid eigenvector are either known pectin metabolisers or species from clades known to include pectin metabolisers, such as *Bacteroides thetaiotamicron*, *Gemmiger qubicalis*, and several *Lachnospira spp*[35–37].

Using univariate tests, we identified two genera that were significantly increased in faecal samples after the HB diet, *CAG-*1427 and *Lachnospira* (𝑞 ≤ 0.05, Wilcoxon signed rank test). *CAG-1427* is part of the Eggerthellaceae family, in our data it was strongly correlated to the carotenoid lutein. Genus *CAG-1427* was present in 90% of our cohort, and represented by 7 species, that were all increased in HB and all unnamed so far. *Lachnospira* was represented by 7 species in our samples, which showed a similar distribution pattern to *CAG-1427,* being broadly represented among samples (e.g. *L. sp003451515* was found in all 19 of 20 participants and 92% of samples), but no species was nominally significant, although all but *L. sp000436475* were increased in relative abundance, supporting the findings from our dbRDA (Figure 2B).

### Functional composition of community reflects taxonomic composition

The functional composition, as represented through KEGG orthologs, showed similar patterns to taxonomic composition. Most (80%) of the functional variance could be explained by interindividual differences (𝑝 ≤ 0.001, PERMANOVA), and diet explaining 2% (𝑝 = 0.28, PERMANOVA). The functional composition after HB diets was significantly more similar between individuals than in either baseline or the LB arm (Figure 2D), and functional diversity was increased in HB (Figure 2C), as also found for taxonomic composition. The functional and taxonomic spaces were highly correlated (Mantel 𝑟 = 0.56, 𝑝 ≤ 0.001).

To better understand the implication of functional changes with either diet, we relied on GMM (gut-specific metabolic modules)[38] to capture important pathways for gut health.

Valine degradation and anaerobic fatty acid beta-oxidation were higher in relative abundance in HB compared to LB (𝑞 < 0.1, paired Wilcoxon rank sum test, BH adjustment). Further, we found significant correlations between several of these pathways and bioactive intake: intake of the carotenoids lycopene and beta-carotene were both positively correlated with anaerobic fatty acid beta-oxidation, glycerol degradation via the propanediol pathway, and dissimilatory sulfate reduction (𝑞 < 0.1, Spearman correlation). Lycopene intake also had a significant positive correlation with the galactonate degradation module, and beta-carotene a negative correlation with the acetate to acetyl-CoA module. Glucosinolate intake also showed a significant positive correlation to valine degradation (𝑞 < 0.1). Magnesium was the only nutrient with significant correlations to these functional modules, and was negatively correlated to mannitol and sorbitol degradation.

To explore connections between individual taxa and their metabolic potential, we investigated carbohydrate active enzymes (CAZymes) and their substrate target, found in each MGS. Over 80,000 genes for 308 CAZyme families were identified in our gene catalogue, linking these to our MGS identified 84 CAZyme genes on median per MGS. Evaluating all MGS found associated to either HB or LB in a constrained dbRDA (Table S3), we found more CAZymes targeting four substrates in either group than the remaining MGS (Figure 2E). Enzymes for degradation of plant cell wall fibres arabinan, xylan, and starch were increased in HB and LB associated MGS compared to MGS with no diet association, while beta-mannan was overrepresented in specifically HB-MGS.

### Urine and faecal metabolomics

In addition to the change in protein coding genes in response to the diet, we used untargeted faecal and urinary metabolomics to assess changes in microbial and human metabolism.

### Microbial and host metabolites shows greater response than microbiome composition

A total of 151 urinary and 60 faecal metabolites showed significant differences when compared between HB and LB (Figure 3A) samples, indicating a more dynamic response to changes in diet than the microbial composition. In urine samples, 119 metabolites were increased in HB, while only 32 metabolites were increased during LB diet interventions. Some of these metabolites likely originate from the HB diet: theobromine directly from, or as a secondary metabolite of, cocoa, chocolate, tea and coffee[39]; or o-coumaric acid which is found in a wide range fruits, vegetables and seeds[40]. In faecal samples, overall fewer significantly different metabolites were found (25 HB, 35 LB). The absolute log fold change was again higher for those metabolites showing differences which were enriched in HB than those in LB (mean 0.96 in LB, 2.04 in HB, 𝑝 ≤ 0.001, Wilcoxon rank sum test). Of the identified peaks, most enriched in HB was 2-Hydroxyphenylacetic acid, but also ursodeoxycholic acid, a secondary bile acid typical for high-fat diets was enriched in HB, though fat intake was significantly lower in HB than LB (Supplementary Figure 3). Identified metabolites found to be lower in HB include 2-Hydroxyhexanoic acid and beta-Pseudouridine.

**Figure 3:**
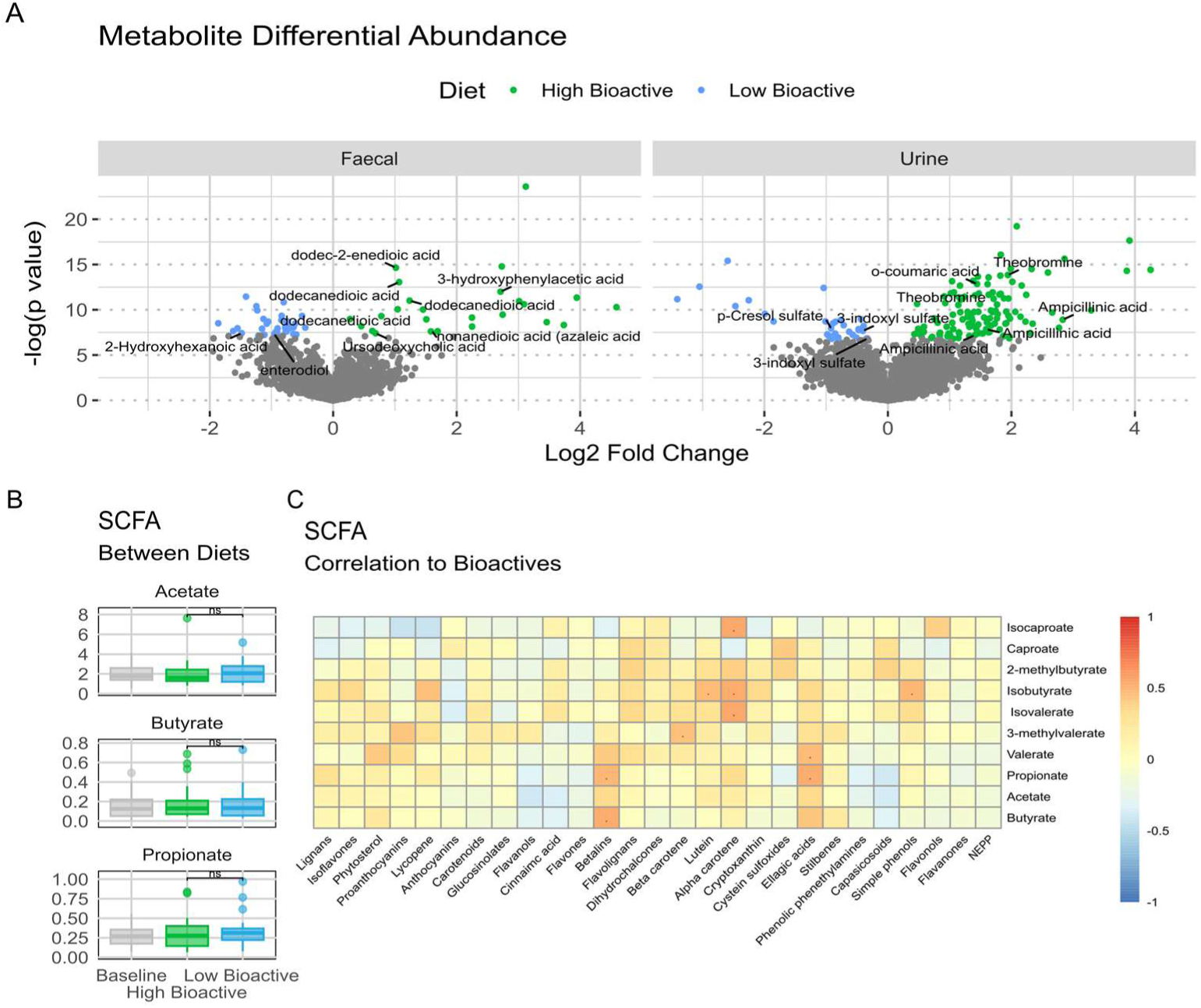
Differences in untargeted metabolomics profiles between HB and LB. A) Change in abundance of features from untargeted metabolomics. Faecal (left) and urine (right) metabolomics were compared between HB and LB samples. Significance tested with a linear model, any metabolite with q <= 0.05 are shown in green if more abundant in HB, blue in LB. Labels have been added for metabolites which have been identified. B) Concentration of short-chain fatty acids (SCFA) in different dietary conditions. C) Pearson correlation between intake of bioactives and SCFA concentration during HB (. p <= 0.05, all q > 0.1 after BH adjustment.)

Overall metabolite profiles were found to be different between BLN, LB, and HB for both faeces and urine, accounting for interindividual variation (p=0.012 faeces, p=0.029 urine, Type III PERMANOVA on Euclidean distance). Metabolite profiles, conditioned for participant identity, further demonstrated that interindividual variation accounted for 43% and 46% of faecal and urinary metabolite variance, respectively.

### Metabolite changes corresponding to types of food consumed

Next, we wanted to explore how bioactive intake impacts faecal metabolites. The concentration of short-chain fatty acids (SCFAs) did not change between LB and HB interventions (Figure 3B). However, during the HB intervention, several SCFA correlated to the intake of specific bioactives (Figure 3C): acetate, propionate, and butyrate, that constitute 80% of all SCFAs, were positively correlated with intake of betalains and ellagic acids intake. SCFA isoforms, such as Isobutyrate, isovalerate and isocaproate correlated to alphacarotene intake, which might indicate an important role of this bioactive in SCFA metabolism, in addition to the documented increase in SCFAs from other carotenoids (beta-carotene, lutein, lycopene, astaxanthin)[41].

To look broadly at the connection between gut microbial community composition and metabolic activity, we performed a dbRDA of Enterosignature (ES) weight conditioned for participants, and fit metabolite concentrations to the first two axes (Supplementary Figure 8). The first two axes are driven by the three ES, which are typically high in abundance in healthy adult samples (ES_Prev, ES_Firm, and ES_Bact). However, more of the significantly correlated metabolites (𝑝 ≤ 0.05) are associated with ES_Bact (46%) than ES_Firm or ES_Prev (23%, 8%, respectively). Among the peaks which were assigned identities, ES_Bact is positively related to 2-Hydroxyhexanoic acid and β-Pseudouridine, and ES_Firm is positively associated to ursodeoxycholic acid and 2-Hydroxyphenylacetic acid (Supplementary Figure 8). The centroids of the three other Enterosignatures (ES_Prev, ES_Esch, ES_Bifi) do not display similarly strong associations to metabolites, indicating the importance of the taxa represented by ES_Bact and ES_Firm in shaping faecal microbial activity as well as composition.

### Hub metabolites in HB associated to transcription and regulation genes

To investigate interrelations between metabolites and the genes producing them, we constructed network models combining abundance of Pfam domains and significantly different faecal metabolites using SPIEC-EASI[42] (Figure 4).

**Figure 4:**
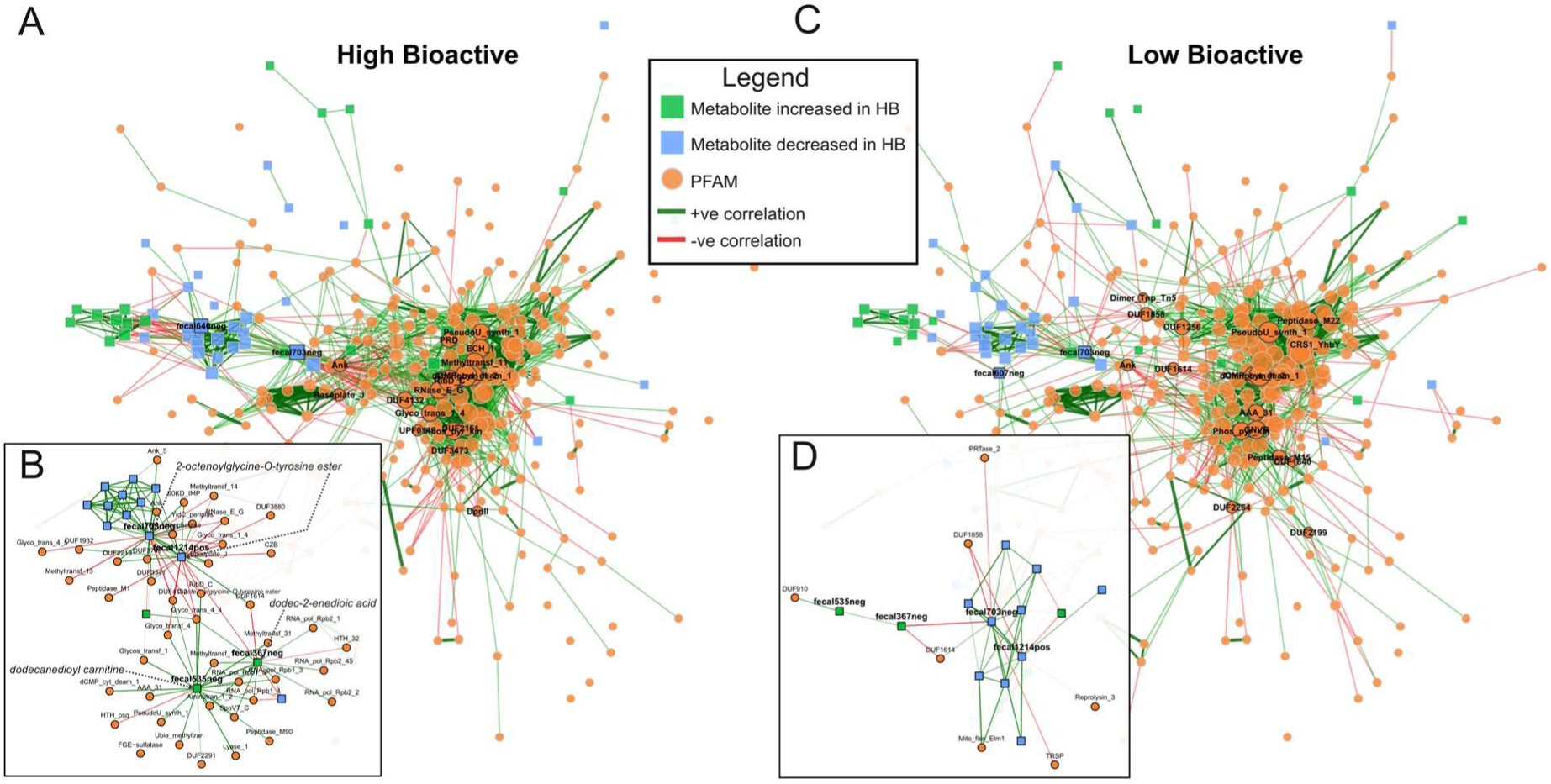
Visualisation of microbiome-metabolite networks, using Pfam composition and selected faecal metabolites. Graphs were constructed separately for HB (A, B) and LB (C, D) samples. Only metabolites were included that are significantly enriched in either HB or LB, respectively. Squares represent metabolites, and circles Pfam domains. Metabolites in green are more abundant in HB, and those in blue more abundant in LB. Vertices with no edges in either graph have been removed. Edges indicate a relationship between two vertices, with colour indicating direction (green positive, red negative), thickness the strength of estimated correlations. Hub vertices based on betweenness are labelled and given a grey border. Inset panels B) and D) show the neighbourhood of four metabolites which are highly connected in the HB graph, and the neighbourhood of those same vertices in the LB graph, reduced to only show Pfam to metabolite and metabolite to metabolite edges.

The HB (Figure 4A) graph shows almost twice as many edges representing relationships between microbial function and metabolites than LB (Figure 4C, 120 to 64 respectively), despite similar overall proportion of possible relationships being identified (graph density 0.025, 0.026 in HB, LB). Edges between Pfams and metabolites in HB are concentrated around a small number of metabolites, with 4 metabolites accounting for 58% of these edges. These four highly connected metabolites and the Pfam domains associated to them can be broadly divided into two communities, with community HBcom-up containing fecal535neg (tentatively identified as dodecanedioyl carnitine) and fecal367neg (identified as dodec-2-enedioic acid) which are more abundant in HB, and HBcom-down containing fecal703neg and fecal1214pos (both are tentatively identified as 2-octenoylglycine-O-tyrosine ester) which are less abundant in HB (Figure 4B).

The two metabolites in HBcom-up are positively correlated with a range of transcription and regulation related domains: they share associations with RNA polymerase Rpb1 domains and methyltransferase domains; fecal367neg is additionally correlated to Rpb2 domains.

SpoVT_C is a domain of the transcription factor involved in bacterial endospore formation, and associated to both metabolites. Domains related to uridine and pseudouridine synthesis (dCMP_cyt_deam_1, CMP_cyt_deam_1) and riboflavin synthesis (RibD_C) are associated to fecal535neg.

The two metabolites in HBcom-down had negative correlations to glycosyltransferase domains and RibD_C, contrasting with the positive correlations between HBcom-up metabolites and these functions. Functions with positive edges to the HBcom-down metabolites include 3 DUF domains among which is DUF3749 that has been linked to acetyltransferase activity[43], as well as membrane related YidC domains (60KD_IMP and YidC_periplas) and ankyrin repeats (Ank_5, Ank). In contrast to HBcom-up, HBcom-down has negative correlations to glycosyltransferases and methyltransferases, as well as to ribonuclease E/G which is involved in cellular RNA degradation and rRNA/tRNA maturation. The associations we found between metabolites and abundance of functions included many domains that could reflect short term changes in activity in response to changes in conditions between HB and LB. Similarly, the increased association to sporulation related domain SpoVT_C in HB is a potential alternative strategy to survive the change in condition driven by differences in diet.

### Diet explains more variance in metabolite profiles than in microbiome composition

The three dietary conditions (BLN, HB, LB) explain 1.13% of variance in metagenomic species composition of participants microbiome, and 5.6% and 5.4% of urinary and faecal metabolite profiles (assessed using dbRDA). Bioactive dietary intake showed no significant correlation to microbiome composition (𝑝 = 0.217, 𝑟 = 0.055 mantel test), while nutrient intake has a significant positive correlation (𝑝 = 0.001, 𝑟 = 0.23, mantel test). Differences in nutrient intake showed stronger correlation to urinary (𝑞 = 0.033, 𝑟 = 0.21, mantel test) than to faecal (p=0.79. r-0.085, mantel test) metabolites; while bioactive intake differences were not correlated to either metabolite measures. This limited impact on overall microbiome composition is reflected in beta diversity between the arms; changes in an individual’s microbiome between dietary conditions show no significant difference (measured with Bray-Curtis dissimilarity, Supplementary Figure 9).

## Discussion

The present study aimed to assess the effect of diets rich in dietary bioactives on healthy subjects’ gut microbial diversity and function using a randomised two period two treatment crossover design. Twenty participants completed the study, each consuming a diet high in bioactive-rich foods for two weeks and, following a washout period, a diet low in bioactive-rich foods for two weeks. The dietary intervention for each of the arms was carefully designed to deliver either high or low levels of dietary bioactive compounds from three key groups, polyphenols, glucosinolates, and carotenoids, but maintaining similar levels of fibre, a key modulator of gut microbiota. This allowed us for the first time to assess the effects of dietary bioactives separate to fibre, which is not usually possible in interventions aiming to increase fruit and vegetables, which inadvertently deliver increase in both.

The design of the study successfully increased participants intake of bioactives from 1.07g at baseline to 4.97g during the HB diet intervention, while in the LB intervention the intake was halved to 0.58 g over a two-week period. Although the total bioactive intake has not been reported before, mean total polyphenol intake ranges between 1.07 and 1.42 g in healthy adults in the UK[44], suggesting that our intervention was effective in significantly and meaningfully altering the levels of bioactives in the diets of the participants. The baseline period represents the composition of the participants typical diets, and bioactive intake was low during baseline, and ordinations of bioactive composition place these baseline diets close to those of the low bioactive intervention.

Dietary advice given to participants to increase or decrease total bioactive intake, depending on the allocated arm, also included advice to achieve the recommended daily fibre intake, which for the UK is 30g[45]. Study subjects managed to increase their total fibre intake to between 26-29g daily in both LB and HB from 22g at baseline. This increase was achieved by carefully selecting foods and included nuts. Increased intake of nuts during the LB dietary arm to maintain constant fibre contributed to a higher total fat intake compared to LB, including higher mono- and poly-unsaturated fats and omega 6 intakes. While a comparable total dietary fibre intake between LB and HB was achieved, the types of fibre that these two interventions delivered will differ, such as increased pectin in HB with more fruit intake, or lignin, cellulose and hemicellulose in LB from increased nut intake, which may affect microbial composition differently[46].

We found small increases in alpha diversity after the HB diet when compared to LB, with diversity of both taxa and encoded microbial functions increasing (p=0.040 and p=0.44 respectively). Previously, increasing ‘healthfulness’ of diet, as assessed by multiple diet quality scores, was strongly associated positively with faecal microbial alpha diversity[47]. Our results suggest that a part of this is driven by the non-nutrient components of ‘healthy’ diets, such as dietary bioactives. While the gut microbiome composition is highly variable between individuals, decreased taxonomic and genetic alpha diversity is often associated with disease[48]. Decreased microbial diversity, such as that seen in gut dysbiosis, has been linked to a variety of non-communicable chronic diseases (NCCDs), including obesity[49], inflammatory bowel disease[50], coeliac disease[51], Type 2 diabetes mellitus[52], certain cancers[53] and general inflammatory processes which may promote cardiovascular disorders[54]. However diversity can be affected by non-disease factors such as transit time and diet[55].

The abundance of two bacterial genera, *CAG-1427* and *Lachnospira*, were also observed to be increased in the HB diet. *CAG-1427* is in the Eggerthellaceae family, species of which have been associated with metabolism of ellagic acid (a polyphenol typically high in berries)[56], and *CAG-1427* was previously found enriched in faecal metagenomes during a high-polyphenol intervention diet[57], and contains a gene cluster for production of the isoflavonoid metabolite equol[57,58] suggesting a direct link to the polyphenol content of the HB diet. Members of genus *Lachnospira* are known to be involved in degradation of dietary fibre pectin found in a range of fruits and vegetables[37,59].

Despite modest microbial taxonomic shifts in response to a two-week high-bioactive diet, overall, the strongest effect of increased bioactive dietary intake was observed in urinary metabolite profiles, followed by the faecal metabolites (for metabolites 5.6% urine and 4.6% faecal variance explained; compared to 1.13% for microbiome composition). Host factors such as age or gender can impact urinary metabolite profiles[60], as well as host processing transforming food derived metabolites to a variety of compounds subsequently detectable in urine[61]. In contrast, the faecal metabolome is mostly the product of microbial metabolism and thus less influenced by the host[62]. Thus, it was important to observe that our diet intervention, matched in calories and nutrients but modified in dietary bioactives, affected faecal metabolite profiles, but only a small number of changes in abundance of individual taxa in the faecal microbiome were observed. This suggests that in the short intervention period with a cohort of 20 participants the functional activity of the community was altered in response to a diet increased in dietary bioactives, while few shifts were detected in bacterial species abundance.

This is relevant, as dietary fibres are known to profoundly shift microbiome composition[63–65]; dietary bioactives seem to rather change the activity of an existing microbial community. This is reflected in our network analysis, which shows associations between domains involved in regulation and transcription and some keystone metabolites whose abundance increases in HB. Further, the host age could also play a role: our subjects had a mean age of 41±20 years old, all being healthy, and their gut microbiome may have been resilient to interventions, having the functional redundancy encoded in their microbiome[66] that allowed a dynamic response to changing bioactive levels.

Both LB and HB diets led to similar total fibre intakes and concentration of SCFAs did not differ significantly between either intervention. However, in our whole-food intervention, several SCFAs significantly correlated to the intake of specific dietary bioactives, such as betalains and ellagic acids. This is in line with existing evidence for an increase of SCFAs and SCFA-producing bacteria in response to single food interventions with red beetroot juice, rich in betalains[67], and strawberries, rich in polyphenols including ellagic acid[68]. In our study genomes recovered for taxa associated to either diet (HB or LB) encoded increased numbers of some plant cell wall fibre degrading enzymes compared to those not associated with either diet. This suggests that in addition to the total dietary fibre amount, the composition of fibre components may also be relevant. Different plant fibre degrading taxa were affected by the increased fibre in the two groups, but the overall output of these taxa was similar between the diets, based on metabolome SCFA measurements. It is possible that the types of dietary fibre such as pectin from increased fruit consumption in the high bioactive diet (Supplementary Figure 6) drive the response of different fibre degrading microbes between the two diets.

## Conclusion

In this study we explored the relationship between dietary bioactives and gut microbes by measuring alterations in faecal and urine metabolites, and gut microbiome composition during a dietary intervention designed to increase dietary bioactives while maintaining similar total fibre levels. We found that dietary bioactives have a relatively small impact on the gut microbiome composition, increasing taxonomic and functional diversity, but greater impact in shaping the metabolic activity of both bacteria and the human host; particularly a high-bioactive diet led to substantial shifts in the metabolic landscape. Genome resolved metagenomics in combination with metabolite-taxonomic co-occurrence networks allowed us to track some of the shifted metabolites to specific functions encoded by bacteria. This dietary intervention study provides insights into how diets rich in fruits and vegetables affect the gut microbiota and highlights the complex interactions between food components that are at play, which strongly supports the careful assessment of dietary intake in all gut microbiome studies. While the total fibre intake was constant between diets, we found some evidence that the types of fibre consumed affect the microbial composition, highlighting the benefit for future study design to consider not just overall fibre but fibre composition, and of work to assess fibre composition of plants based foods. In addition, the impact of this study goes beyond the dietary field, demonstrating the health impact certain foods can have by altering bacterial and host metabolite profiles. This reinforces the need for a paradigm shift in the gut microbiome field, for studies to move beyond the “composition only” approach, but rather focus (or at least include) bacterial activity when investigating microbiomes.

## Materials and methods

### Study design

This was a nine-week randomised, cross-over, pilot study where all visits were conducted remotely due to the national lockdown that was imposed because of the COVID-19 pandemic in 2020 (Figure 8). The study began with a 1-week run-in (baseline) where participants provided a 7-day food diary about their habitual diet and applied a continuous glucose monitor (CGM) (Freestyle Libre2 from Abbott Diabetes Care, Alameda, USA), see Figure 1A. After the run-in period, participants were randomised, and intervention commenced following a 2-hour oral glucose tolerance test, which was then followed by a 2-week intervention with one of the two diets (high or low bioactive). After completion of the first intervention, participants then had crossed over to the alternate diet. A 4-week washout separated the two intervention periods. Within each intervention, an oral glucose tolerance test was nested before and at the end of the intervention period. Additionally, participants were asked in the three days leading to the end of the intervention period to consume a standardised breakfast for three days. Participants were asked to wear a continuous glucose monitor (CGM) and a Fitbit Inspire HR during the run-in and intervention periods. The CGM monitored the postprandial changes in interstitial glucose after the glucose beverage and standardised breakfasts. 24-hour urine and faecal samples were collected before each intervention period as well as one extra faecal sample at midpoint of the washout period. Each remote visit was conducted using a virtual meeting platform (Microsoft Teams) and began with weight, waist circumference, blood pressure, and pulse measures.

### Participants

Participants were recruited within 40 miles of the Norwich Research Park, Norwich, UK. Eligible participants were 18–65 years old with a body mass index (BMI) between 18.5 and 35 kg/m2, HbA1c < 42 mmoL/L, had access to a smart mobile phone with near field communication (NFC) technology, and resided in an area where online food deliveries could be made. Participants were excluded if they were smokers; had abnormal blood pressure (<90/50 or ≥160/100 mmHg); had any allergies to foods that were included in the dietary intervention; had any conditions or on medication that could alter metabolism (i.e. diabetes mellitus, hypothyroidism, etc.); have altered absorption or digestion (ulcerative colitis, histamine blocking medications, etc); have altered immune function (anaemia, lupus, chemotherapy, etc), have cardiovascular disease; undergoing treatment for cancer; used antibiotic 6 months prior to study; frequently undergoes bowel cleansing procedures; or unable to discontinue dietary or probiotic supplements. Women who were pregnant, lactating, or had given birth in the last 12 months were also excluded. Written and verbal informed consent was obtained and witnessed during a virtual meeting from each enrolled participant.

### Sample Size Estimation

Preliminary estimates of the effect of a high-bioactive diet vs a low-bioactive diet on gut microbiota diversity and composition for this pilot study was based on previous studies that observed differences between groups with different diets between 0.2 and 0.4[11,69] found that Shannon index had a within-person standard deviation of approximately 0.21, which would imply that the standard deviation of difference between the Shannon index on two occasions within a person is √2 × 0.21 = 0.29. Using this value, our expected standard error for the difference in Shannon index based on 20 individuals completing the study would be 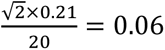. Hence in terms of previously observed effects of diet on Shannon index this study will provide a reasonable preliminary estimate of the effect of bioactive diet on metagenomic diversity, on which further research can build.

### Intervention Diet and Dietary Adherence

Participants were instructed to record their dietary intake of their habitual diet during the 7-day baseline period and for 14 days during each intervention period. Participants when randomised to the high bioactive diet were instructed to consume abundant amounts of pigmented fruit and vegetables into their diet as well as 100 g of mixed summer fruits and 100 mL elderberry juice (Biona, Surrey, UK) daily. Participants when instructed to follow a low bioactive diet participants were asked to avoid pigmented fruits and vegetables as well as drink 100mL unsweetened pineapple juice. For both high and low bioactives diet participants were encouraged to maintain their dietary intake of fibre between 27 to 33g/day. Participants were provided food ingredients, meals, sample menus and recipes to help reduce their burden in changing their diets. Participants recorded their daily energy and nutrient intake using Libro mobile app (Nutritics, Dublin, Ireland).

### Standardised Breakfast

The 3 mornings leading up to the last day of an intervention period, participants were asked to consume a standardised breakfast at the same time each morning. The first standardised breakfast was 2 white bread rolls (Warburtons sliced white rolls, Bolton, UK). Each bread roll was 55g and provided 25g total carbohydrates. The second morning breakfast was 2 white rolls and two (6.2 g each) foil portions of butter (Lakeland Dairies Salted Butter, Cavan, IE).

The third breakfast was 2 white rolls, two portions of butter, and 160g chicken breast slices (Sainsbury’s Ready to Eat Chargrilled, Holborn, UK)

### Metagenomic DNA sequencing preparation

DNA samples were extracted using the Maxwell RSC PureFood and GMO Authentication Kit (Promega #: AS1600) according to the manufacturing instructions. Briefly, 1 mL of CTAB buffer was transferred to 200 mg of faecal sample and vortexed for 30s. Samples were incubated at 95 degrees for 5 minutes and then vortexed for 1 minute. Samples were then homogenised using the fast prep for 45 seconds at 6.0 m/s. Following homogenisation, 40 μL of proteinase K and 20 μL of RNase A were added and briefly vortexed, following 10-minute incubation at 70 degrees. Samples were centrifuged at 14,000 g for 5 minutes, and 300 μL of supernatant was transferred to well 1 of the cartridge along with 300 μL of lysis buffer.

DNA was then quantified using the Qubit dsDNA quantification broad range kit (ThermoFisher #Q32853). Genomic DNA was normalised to 5 ng/µl with EB (10 mM Tris-HCl). Libraries (100 DIME samples plus two negative control waters) were prepared according to Ravi and colleagues[70]. The pooled library were then sent to an external provider, (GeneWiz). 50 million reads were generated per each individual library. The sequencing configuration was 2x 150 bp on an Illumina NovaSeq 6000 system through two S4 lanes.

### Targeted analysis of short-chain fatty acids

Faecal samples were extracted and processed for untargeted metabolomics and for SCFA measurements as described by Pekmez and colleagues[71]. Samples were analysed by ultra-performance liquid chromatography (UPLC) coupled with a quadrupole-TOF mass spectrometer (q-TOF-MS) equipped with ESI (Waters, Manchester, UK). Reverse phase HSS T3 C18 column (2.1 × 100 mm, 1.8 μm) coupled with a pre-column (VanGuard HSS T3 C18 column, 2.1 × 5 mm, 1.8 μm) was used for chromatographic separation.

Five µL of each sample and of a sample pool all containing isotopically labelled internal standards of all targets were injected into the gradient mobile phase A (0.01 % formic acid in Milli–Q water) and mobile phase B (0.01 % formic acid in Acetonitrile). The SCFA intensities were preprocessed in MZmine version 2.28. The internal standard corrected responses, and the quantification of SCFAs were calculated using an in-house script in MATLAB R2015b. If the response for a given analyte was showing a concentration in feces below 0.195 µM, the concentration is reported as under the limit of quantification (ULQ).

### Cell counts

For the flow cytometric enumeration of microbial cells, 100 mg of the faecal samples were dissolved in 0.85 % NaCl (Sigma-Aldrich) and filtered using a 40 µM filter. Filtered samples were diluted 1:20,00 and stained with SYTO9 11 µM (Thermo Fisher Scientific), incubated at 37 degrees for 1 h upon quantification through the Image Stream (Amnis) flow cytometer. Fluorescence events were monitored using the 485 nm optical detectors. Forward and sideways-scattered light was also collected. Flow cytometry measurements were not available for participant 20 for sample at V1 due to insufficient stool volume.

### Untargeted metabolomics

#### Sample preparation

Urine samples were centrifuged (4°C, 4 min, 3000 RCF) and to each well in 96-well collection plates was added a mixture of 150 μL of a urine sample together with 150 μL of solvent (water with 5% 30:70 (v/v) ACN: MeOH) containing a solution with 7 Internal Standards as described previously[72]. In addition, an external metabolite standard mixture with 44 different compounds was used for quality control of the analytical platform[72]. A pooled sample containing equal amounts of all urine samples analysed was prepared and added to separate wells on all plates for subsequent control of instrumental drift and batch effect in the data preprocessing and analysis. Samples from the same person were randomised within one plate to minimise intra-individual variation due to plate differences. After preparation, the plates were sealed and kept at 4 °C if analysed within the next 24 h; they were otherwise kept at −80°C and thawed and gently agitated prior to analysis.

Faecal samples (25mg) were mixed 1:1 with water and ethanol containing 7 internal standards[72] was added to a final concentration of 92%. The samples were centrifuged at 20,800 RCF for 2 minutes in Eppendorf tubes and supernatants filtered through a 0.22µm filter to remove any particulates. The clean samples were placed on a 96-well plate along with a pool of all samples, a blank sample and a mixture of 44 standards as for urine.

#### LC-MS analysis

Urine and faecal samples were profiled in a 7-min gradient run on the same column and precolumn used for the SCFA. The gradient was as described previously (Barri et al., 2012)[72].

#### Pre-Processing

The raw LC-MS data was converted into .mzmL files using ProteoWizard[73]. The R (https://www.R-project.org) package XCMS[74] was used for pre-processing. The CentWave[75] algorithm was used for peak-picking and peaks were grouped across samples using the peak density method and aligned using features common to most samples and finally missing values were “gap-filled” by re-integrating samples for which values were missing for any of the features. CAMERA[76] was used to group related features, to estimate the number of unique compounds and to aid identification.

Missing values (after gap-filling) were filled by feature-wise substitution with a random number between zero and the minimum detected intensity. Features with a D-ratio[77] greater than 0.5 were removed. Each feature was then first corrected for intra-batch drift by fitting a LOESS regression to the QC samples, then the mean of the QCs was used to correct inter-batch differences. Probabilistic Quotient Normalization (PQN) was then used to correct for overall concentration differences.

#### Identification

Compounds were identified by comparing spectral similarity and retention time to an in-house database of authentic standards. All reported compounds are thus identified at MSI[78] Level 1.

Specifically for the fecal metabolites related to the cluster of highly altered bacterial gene expression, additional identifications were done by investigating mass spectral fragmentation in MS and MS/MS (DDA) modes, peak shapes of extracted ion chromatograms, and comparing series of analogues, e.g. medium-chain fatty dicarboxylic acids and their monounsaturated analogs; for instance C8-C14 dioic acid chain lengths and monounsaturation have a linear relationship with their retention times, showing that the MS/MS spectral interpretation coincide with retention time prediction of the structure of dodecanedioic acid. Both this and a detailed description of the putative identification of 2-octenoylglycine-O-tyrosine ester is provided in Supplementary Results 1.

### Metagenome assembly and recovery of metagenomic species

Sequences were processed with the MG-TK pipeline[79,80]. Briefly, raw shotgun metagenomes were quality filtered using sdm v1.63 with default parameters[81], assembled using megahit v 1.2.9 with parameters “--k-list 25,43,67,87,101,127”[82] and reads mapped onto assemblies using bowtie2 v2.3.4.1 with parameters “--end-to-end “[83], genes predicted with prodigal v2.6.1 with parameters “-p meta”[84] and a gene catalogue clustered at 95% nt identity using mmseqs2[85]. MAGs (metagenomic assembled genomes) were binned using semibin[86] and combined in MG-TK to MGS (metagenomic species), relying on canopy clustering[31]. Matrix operations were carried out using rtk[87]. The full MG-TK pipeline is available at https://github.com/hildebra/mg-tk[88].

MGS phylogenies were calculated de novo based on the amino acid (AA) sequences of at most 40 marker genes[89] present in each MGS and subsequently aligned using MAFFT v v7.464[90] with default options; the multiple sequence alignment (MSA) was then trimmed and translated from nucleotide to amino acid sequences using TrimAl[91] (options “-keepheader -ignorestopcodon -gt 0.1 -cons 60”), from these IQ-TREE v1.6.3.a[92] with parameters “-m GTR+F+I+G4 -B 1000” was used to reconstruct a phylogeny, that was visualised with iTOL[93].

The abundance of these MGS are estimated based on the coverage of their gene content. The abundance of species for which an MGS was not recovered are estimated by the coverage and taxonomic annotation of the genes which do not fall within any of the MGS.

### Statistical and metagenomic analysis

Initial descriptive analysis of participant clinical and dietary measures are reported as mean ± standard error of mean (SEM) (Supplementary Table 1). A student’s t-test was used to make paired comparisons between each period where dietary records were collected (Baseline, HB diet, and LB diet). To adjust for multiple comparisons, a test was considered significant if its p value was less than 0.01 as opposed to the traditional 0.05.

Dietary intake data for each component other than total energy intake was subsequently adjusted based on a regression against total energy, to be mean intake plus residual of regression against total energy[30]. PCA analyses were carried out separately on bioactives (n=27) and nutrients (n=101) using a centred scaled transformation. Beta dispersion and PERMANOVA tests were performed using the betadisper and adonis2 functions from vegan[94] on Euclidean distances between centred scaled data, and participant as blocking factor, with post-hoc tests carried out using adonisplus[95]. Mantel and Procrustes tests comparing bioactive and nutrient dissimilarity matrices used the same parameters, and the mantel and protest functions from vegan[94]. Difference for each nutrient between high and low bioactive arms was tested used paired Wilcoxon signed rank tests.

Alpha diversity was calculated using rtk[96], and the mean of 100 rarefactions to the minimum sample sum was used. Difference between interventions was tested using a paired Wilcoxon signed rank test. Bray-Curtis dissimilarity was calculated between MGS level abundances and difference between interventions tested using Wilcoxon signed rank test.

Differences in abundance at each rank for both taxonomic and functional composition was tested using paired Wilcoxon signed rank tests on relative abundance, with Benjamini & Hochberg adjustment.

CAZymes for all genes were predicted using dbCAN-3[97], and results and substrate annotation from the dbsub tool used for quantifying number of enzymes in MGS active on different carbohydrate substrates. To identify diet associated taxa, MGS level abundance was filtered to only samples from BLN, HB and LB, and a dbRDA was constructed using the capscale function in R package vegan with participant identity as a conditioning term and diet as a constraint, using Bray-Curtis dissimilarity. Diet associated taxa were identified based on species scores, taking the 75^th^ quantile of vector length, and considering any vector with cosine ≥ 0.9 with the diet centroids as associated with that diet.

Mantel tests for correlation between nutrient or bioactive intake and metabolite and microbiome compositions were carried out using the mantel function in R package vegan using pearson correlation and 999 permutations. Nutrient and metabolite distances were Euclidean distance based on matrices centred on 0 and scaled by standard deviation.

Microbiome composition used Bray-Curtis dissimilarity on relative abundance of MGS. Tests were adjusted using Benjamini & Hochberg method.

Difference in metabolites concentrations from untargeted metabolomics between HB and LB were tested with paired t-tests and BH adjustment.

Networks combining functional abundance and untargeted metabolomics were constructed using SPIEC-EASI[42,98] with ‘glasso’ method, ‘bstars’ model selection criterion, 100 random subsamples in pulsar, and lambda.min.ratio of 0.1 and nlambda of 100 as parameters for huge. Networks were subsequently analysed using igraph[99] and NetCoMi[100].

## Declarations

### Ethics approval and consent to participate

The study protocol was approved by the Quadram Institute Human Research Governance Committee and the Department of Research and Development (R&D) of the Norfolk and Norwich University Hospitals NHS Foundation Trust. Favourable ethical opinion was received from the Health Research Authority England (Greater Manchester West Research Ethics Committee, REC#:20/NW/0359). The study was registered with International Standard Randomised Controlled Trial Number (ISRCTN14510401). All participants’ data were stored in accordance with the General Data Protection Regulation 2018. Participants consented to have their personally identifiable information shared with third party shops for delivery of food whereas the remaining third-party vendors were only provided pseudo-anonymised numerical identifier. Biological samples were collected remotely, transported by the study team, and disposed of in accordance with the Human Tissue Act (2004).

## Consent for publication

Not applicable

## Availability of data and material

Metabolomics have been uploaded to Metabolights[101] under accessions MTBLS8100 and MTBLS7290 for untargeted and targeted respectively. Faecal sample sequencing is available on ENA under project accession PRJEB55607. Continuous glucose monitoring and sleep data are available on Zenodo (https://doi.org/10.5281/zenodo.8268744). Analysis scripts and dietary intake data are available at https://github.com/apduncan/dime_analysis.

## Authors’ contributions

Conceptualization: MT, FH; Data curation: DN, AD; Formal analysis: FB, DN, JS, LOD, GS, LZ, DS-L, AD; Investigation: FB, JA-J, JS, LOD; Methodology: MT, FH, GS; Supervision: MT, FH; Visualization: AD, JA-J; Writing – original draft: FB, AD, JS, LOD; Writing – review & editing: FH, MT, JB.

## Data Availability

Metabolomics have been uploaded to Metabolights under accessions MTBLS8100 and MTBLS7290 for untargeted and targeted respectively. Faecal sample sequencing is available on ENA under project accession PRJEB55607. Continuous glucose monitoring and sleep data are available on Zenodo (https://doi.org/10.5281/zenodo.8268744). Analysis scripts and dietary intake data are available at https://github.com/apduncan/dime_analysis.

https://www.ebi.ac.uk/metabolights/editor/study/MTBLS8100/descriptors

https://www.ebi.ac.uk/metabolights/editor/study/MTBLS7290/descriptors

https://www.ebi.ac.uk/ena/browser/view/PRJEB55607

https://doi.org/10.5281/zenodo.8268744

https://github.com/apduncan/dime_analysis

## Acknowledgements

Not applicable

