## Supplementary material for "Dietary bioactives increase gut microbiome diversity and alter host and microbial metabolite profiles"

### Supplementary Figures

#### CONSORT diagram

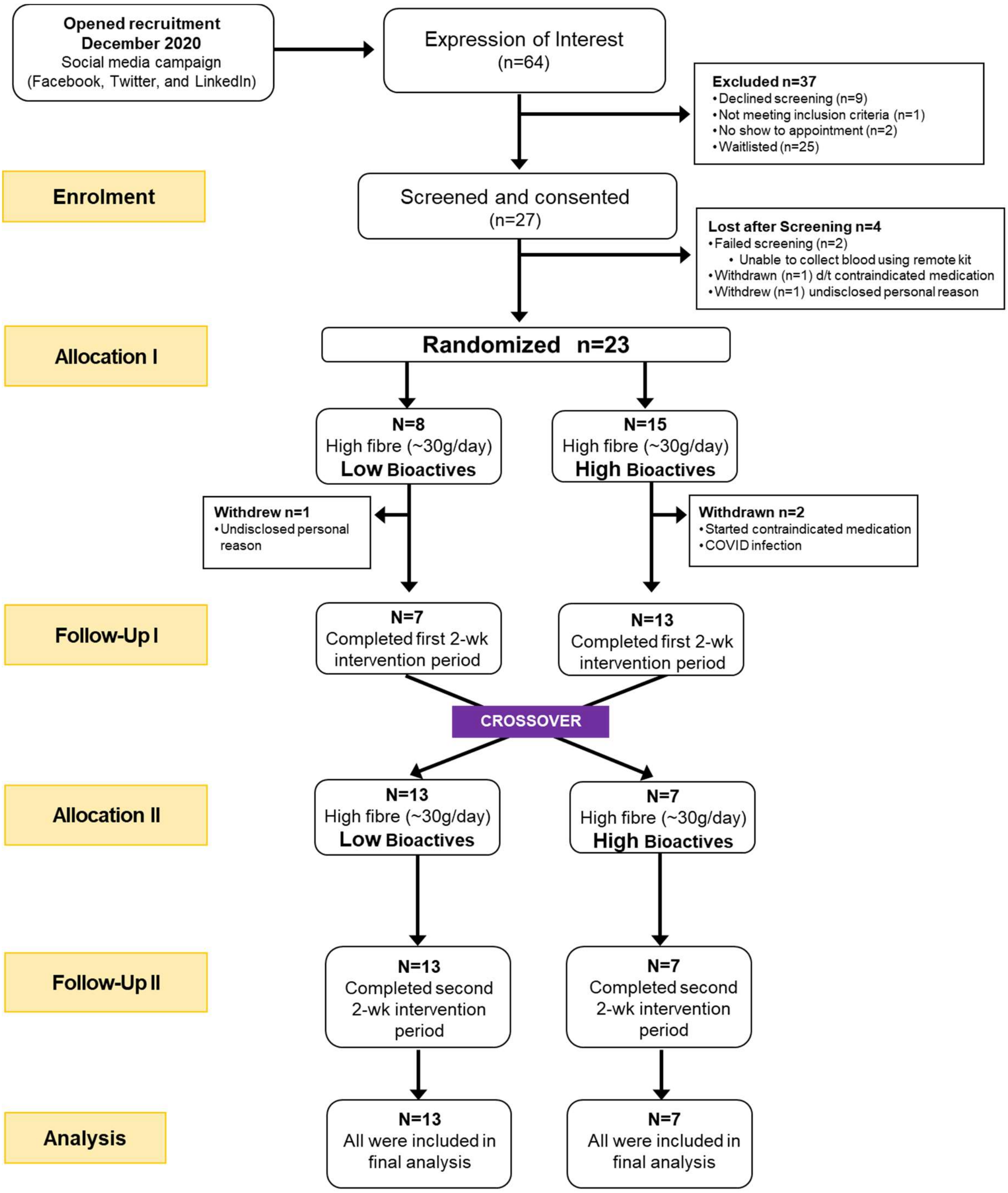


*Figure 1: CONSORT diagram.*

#### Bioactive intake

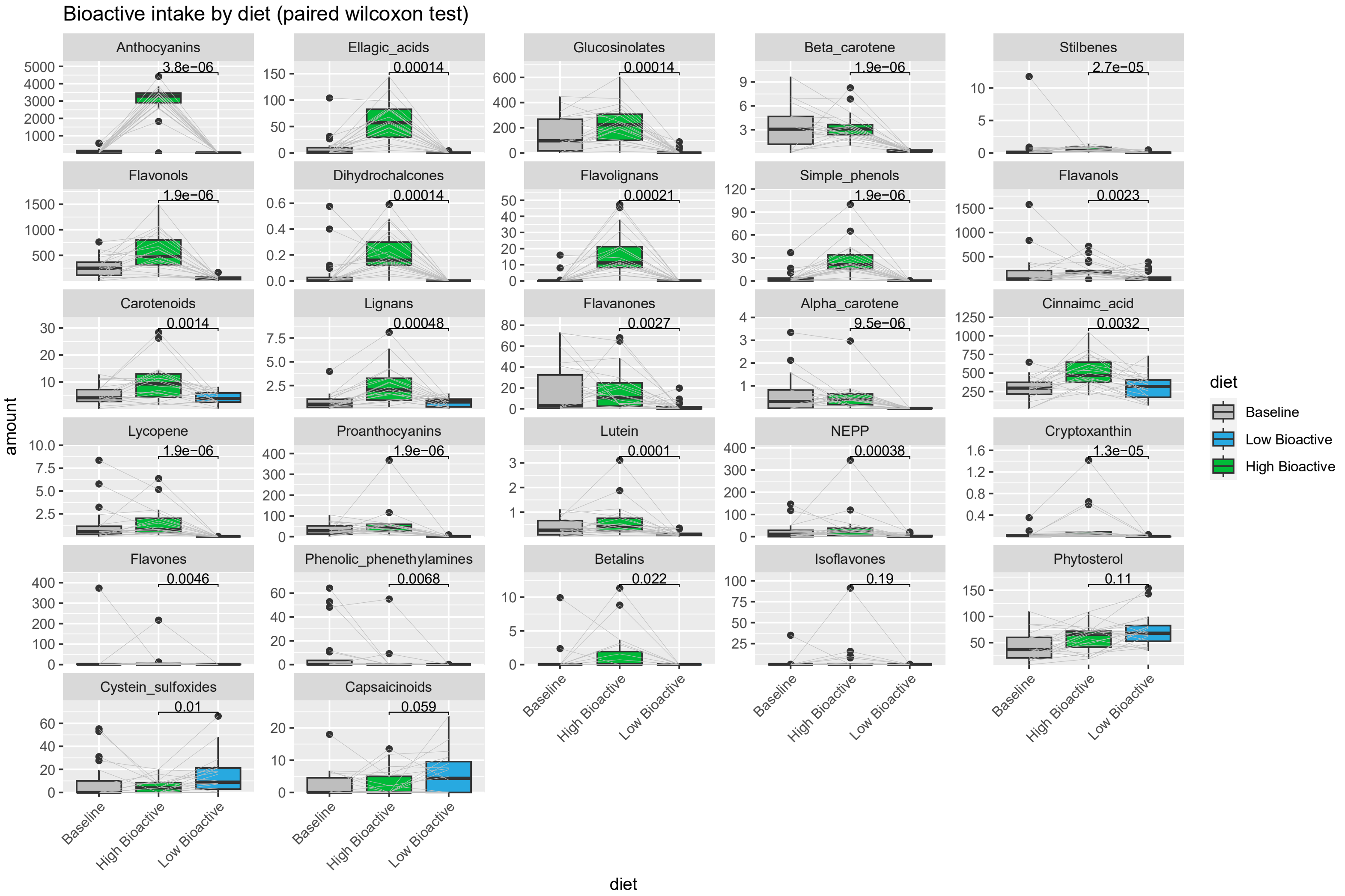


*Figure 2: Intake of bioactives in baseline, high and low bioactive arms of study before adjustment based on regression against total energy intake. Comparison between low and high bioactive is performed using paired Wilcoxon test*

### 9 Nutrient intake

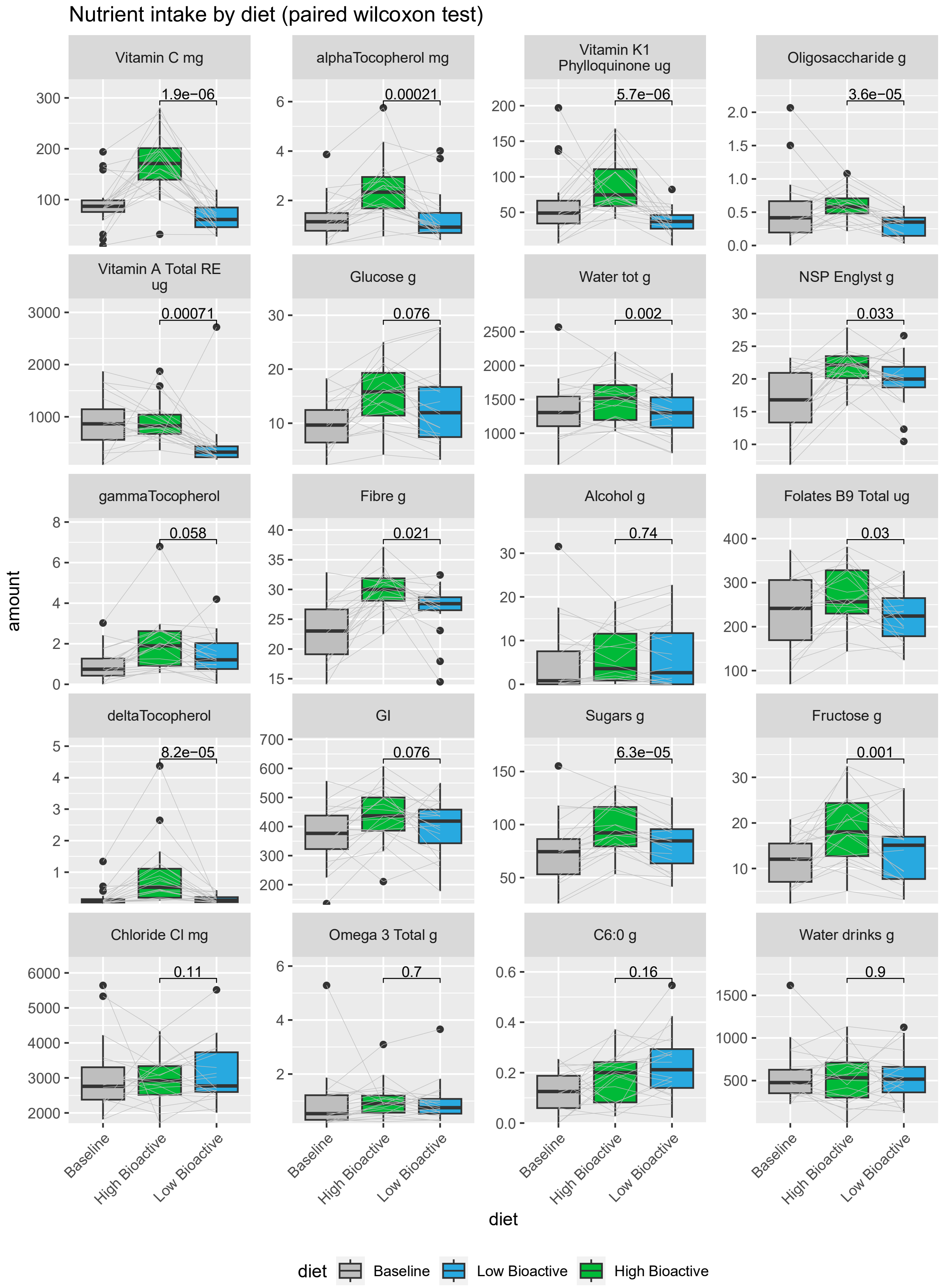

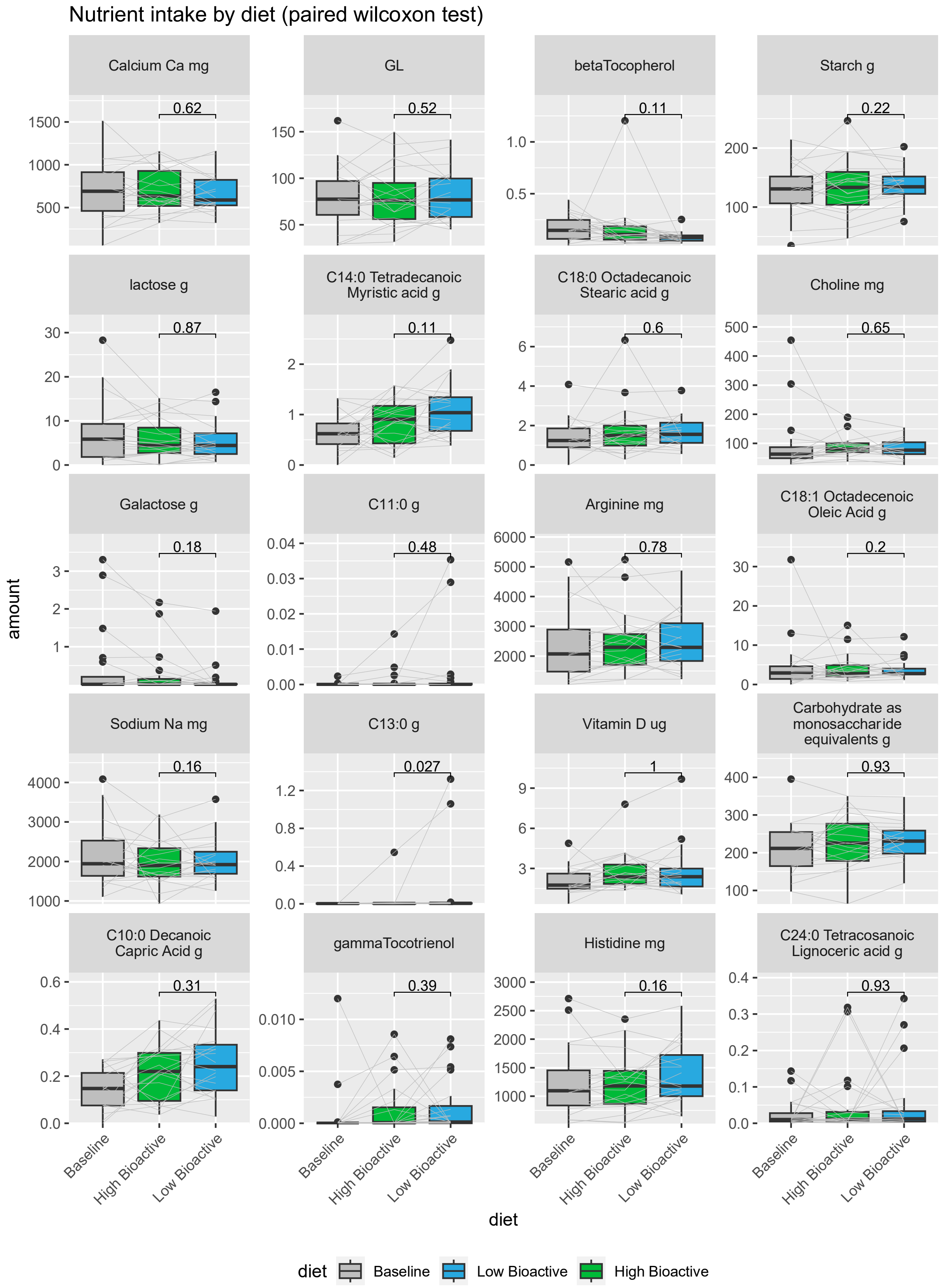

Nutrient intake by diet (paired wilcoxon test)
Calcium Ca mg
0.62
GL
0.52
betaTocopherol
0.11
Starch g
0.22
lactose g
0.87
C14:0 Tetradecanoic Myristic acid g
0.11
C18:0 Octadecanoic Stearic acid g
0.6
Choline mg
0.65
Galactose g
0.18
C11:0 g
0.48
Arginine mg
0.78
C18:1 Octadecenoic Oleic Acid g
0.2
Sodium Na mg
0.16
C13:0 g
0.027
Vitamin D ug
1
Carbohydrate as monosaccharide equivalents g
0.93
C10:0 Decanoic Capric Acid g
0.31
gammaTocotrienol
0.39
Histidine mg
0.16
C24:0 Tetracosanoic Lignoceric acid g
0.93
amount
Baseline
High Bioactive
Low Bioactive
diet
diet
Baseline
Low Bioactive
High Bioactive

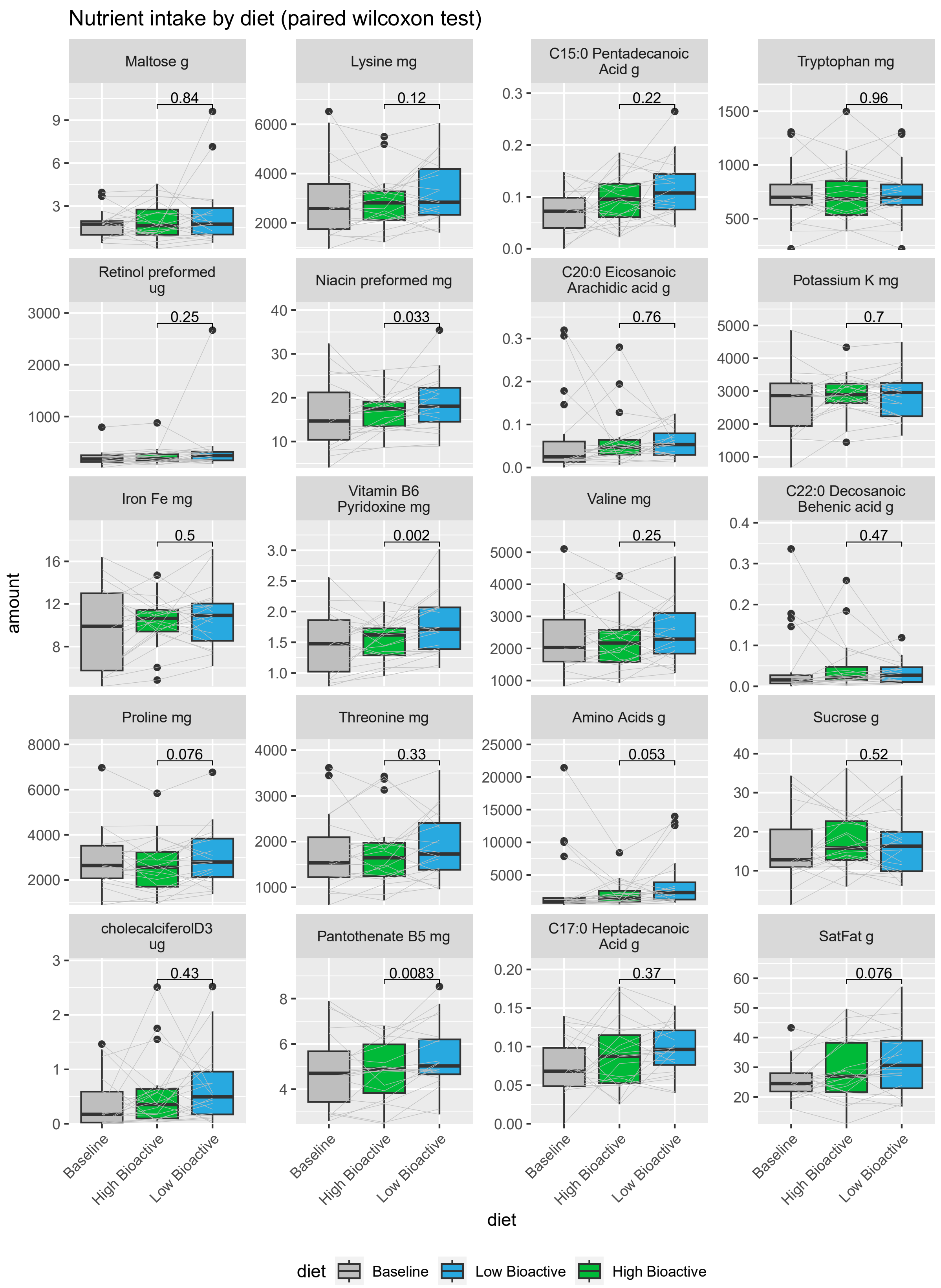
Nutrient intake by diet (paired wilcoxon test)
Maltose g
0.84
Lysine mg
0.12
C15:0 Pentadecanoic Acid g
0.22
Tryptophan mg
0.96
Retinol preformed ug
0.25
Niacin preformed mg
0.033
C20:0 Eicosanoic Arachidic acid g
0.76
Potassium K mg
0.7
Iron Fe mg
0.5
Vitamin B6 Pyridoxine mg
0.002
Valine mg
0.25
C22:0 Decosanoic Behenic acid g
0.47
Proline mg
0.076
Threonine mg
0.33
Amino Acids g
0.053
Sucrose g
0.52
cholecalciferolD3 ug
0.43
Pantothenate B5 mg
0.0083
C17:0 Heptadecanoic Acid g
0.37
SatFat g
0.076
amount
Baseline
High Bioactive
Low Bioactive
diet
diet
Baseline
Low Bioactive
High Bioactive

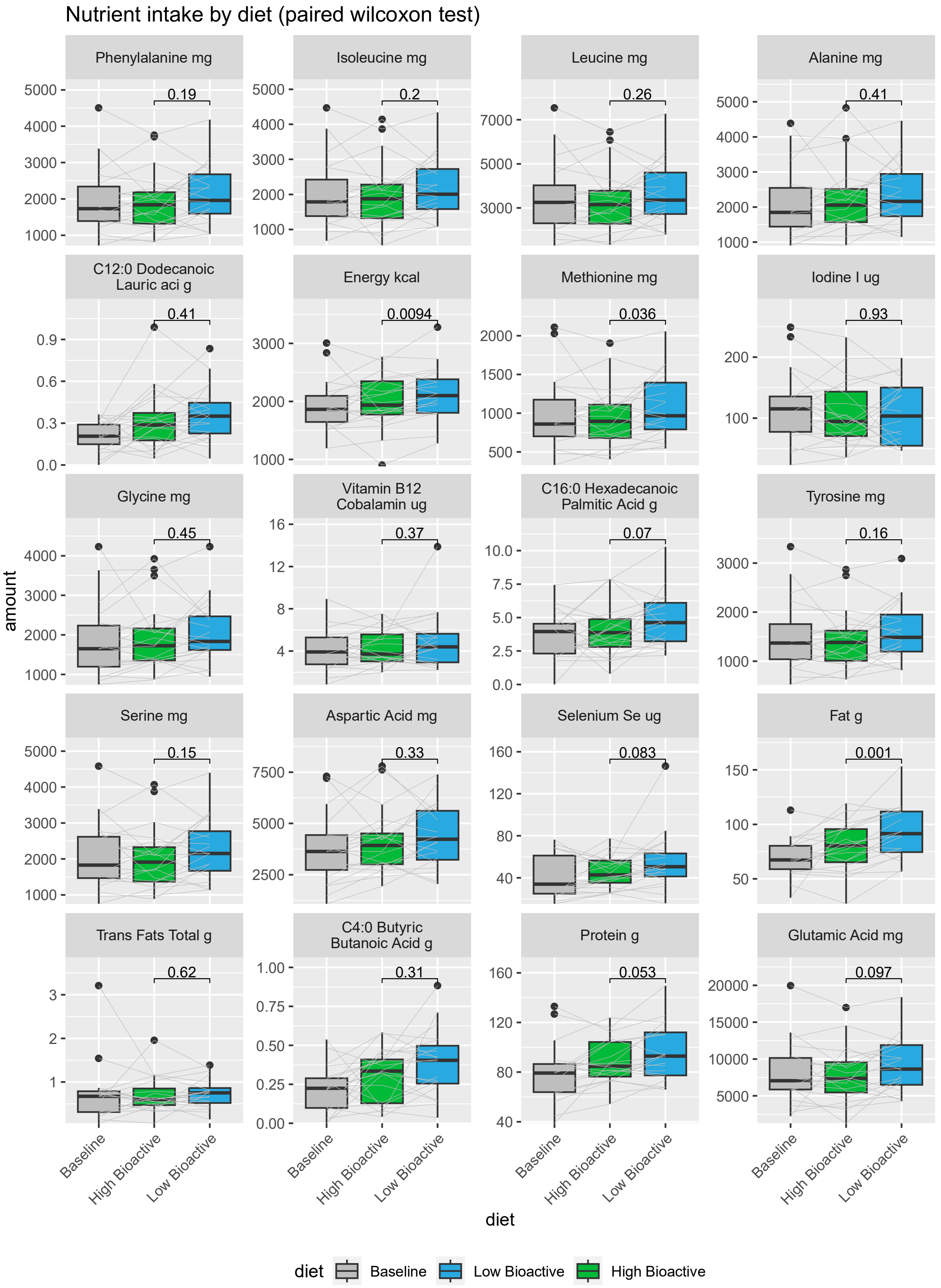
Nutrient intake by diet (paired wilcoxon test)
Phenylalanine mg
0.19
Isoleucine mg
0.2
Leucine mg
0.26
Alanine mg
0.41
C12:0 Dodecanoic Lauric aci g
0.41
Energy kcal
0.0094
Methionine mg
0.036
Iodine I ug
0.93
Glycine mg
0.45
Vitamin B12 Cobalamin ug
0.37
C16:0 Hexadecanoic Palmitic Acid g
0.07
Tyrosine mg
0.16
Serine mg
0.15
Aspartic Acid mg
0.33
Selenium Se ug
0.083
Fat g
0.001
Trans Fats Total g
0.62
C4:0 Butyric Butanoic Acid g
0.31
Protein g
0.053
Glutamic Acid mg
0.097
amount
Baseline
High Bioactive
Low Bioactive
diet
diet
Baseline
Low Bioactive
High Bioactive

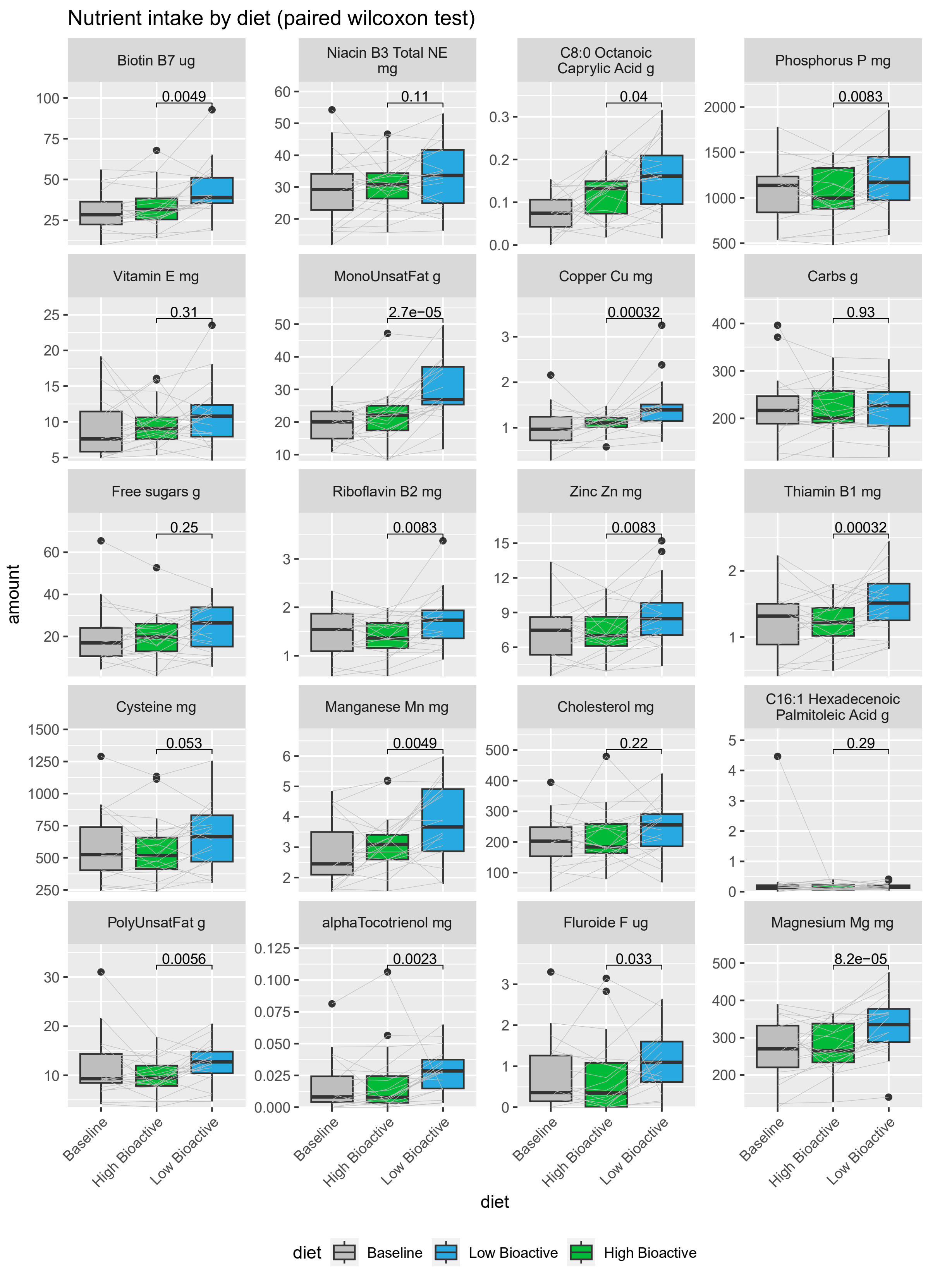

Nutrient intake by diet (paired wilcoxon test)
Biotin B7 ug
0.0049
Niacin B3 Total NE mg
0.11
C8:0 Octanoic Caprylic Acid g
0.04
Phosphorus P mg
0.0083
Vitamin E mg
0.31
MonoUnsatFat g
2.7e−05
Copper Cu mg
0.00032
Carbs g
0.93
Free sugars g
0.25
Riboflavin B2 mg
0.0083
Zinc Zn mg
0.0083
Thiamin B1 mg
0.00032
Cysteine mg
0.053
Manganese Mn mg
0.0049
Cholesterol mg
0.22
C16:1 Hexadecenoic Palmitoleic Acid g
0.29
PolyUnsatFat g
0.0056
alphaTocotrienol mg
0.0023
Fluroide F ug
0.033
Magnesium Mg mg
8.2e−05
amount
Baseline
High Bioactive
Low Bioactive
diet
diet
Baseline
Low Bioactive
High Bioactive

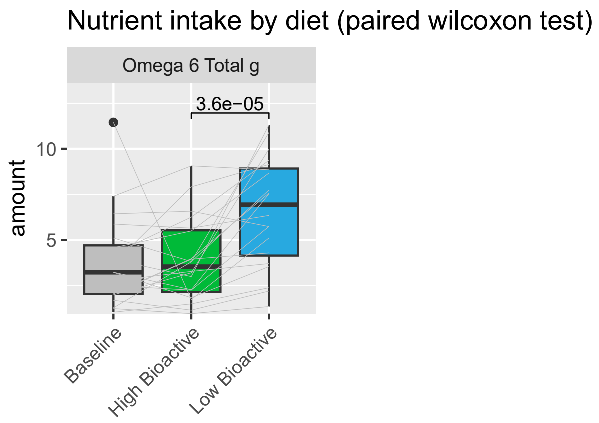


*Figure 3: Intake of nutrients in baseline, high and low bioactive arms of study before adjustment based on regression against total energy intake. Comparison between low and high bioactive is performed using paired Wilcoxon tests.*

#### $R^2$ for bioactives and nutrients regressed against total energy intake

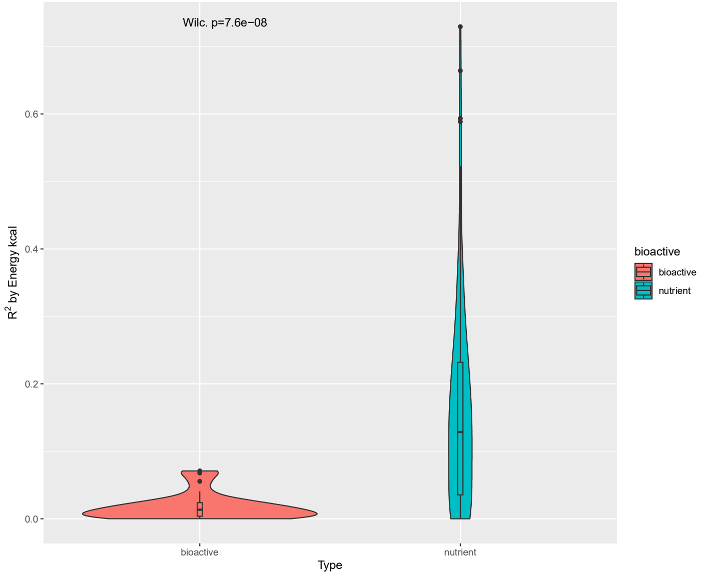


*Figure 4: Distribution of $R^2$ values for linear regression of bioactives and nutrients against total energy intake.*

#### Taxonomic alpha diversity indices

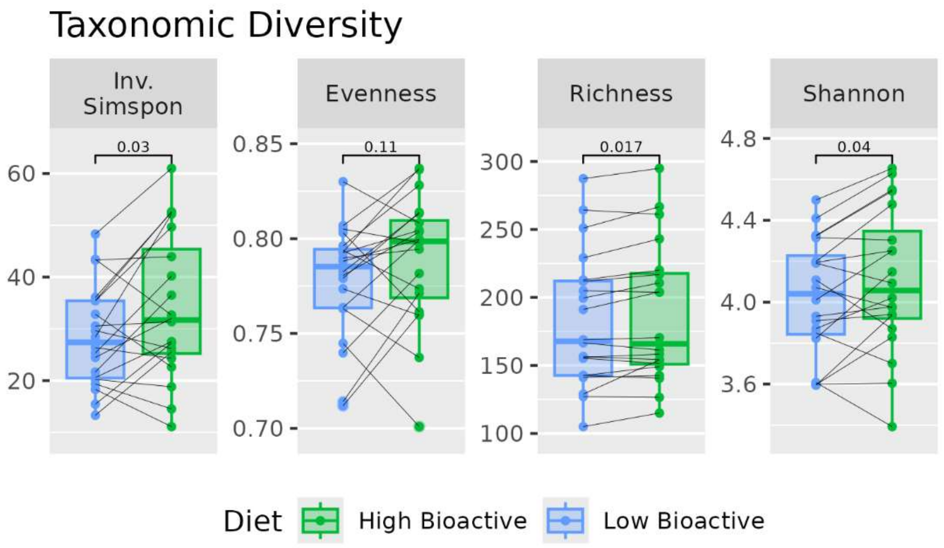


*Figure 5: Measures of taxonomic diversity at species level comparing HB and LB, with significance from Wilcoxon signed rank tests.*

#### Flow cytometry cell counts

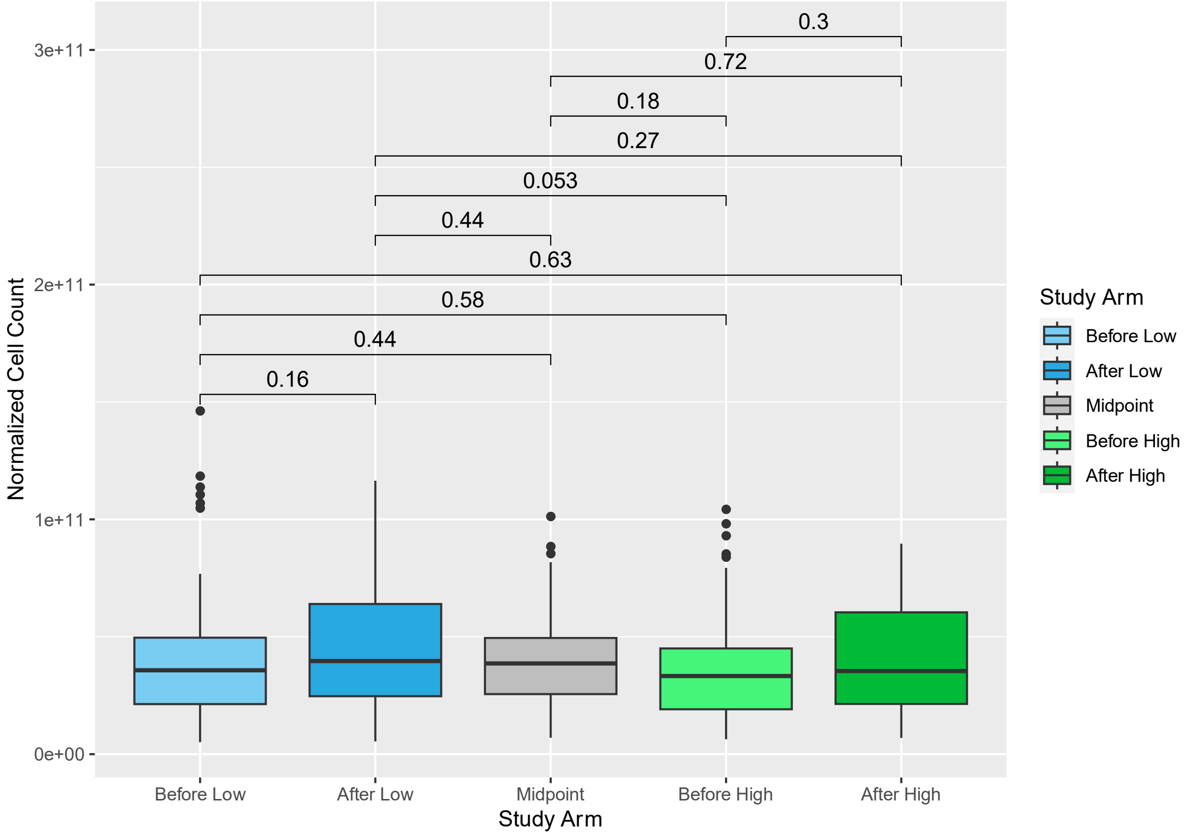


*Figure 6: Distribution of normalized cell counts for flow cytometry experiments from all study arms. Experiments were replicated three times per sample, all replicates included in this plot. Comparison between arms uses Wilcoxon test, without correction for multiple tests. None of the differences between arms are significant.*

#### Enterosignature relative abundance

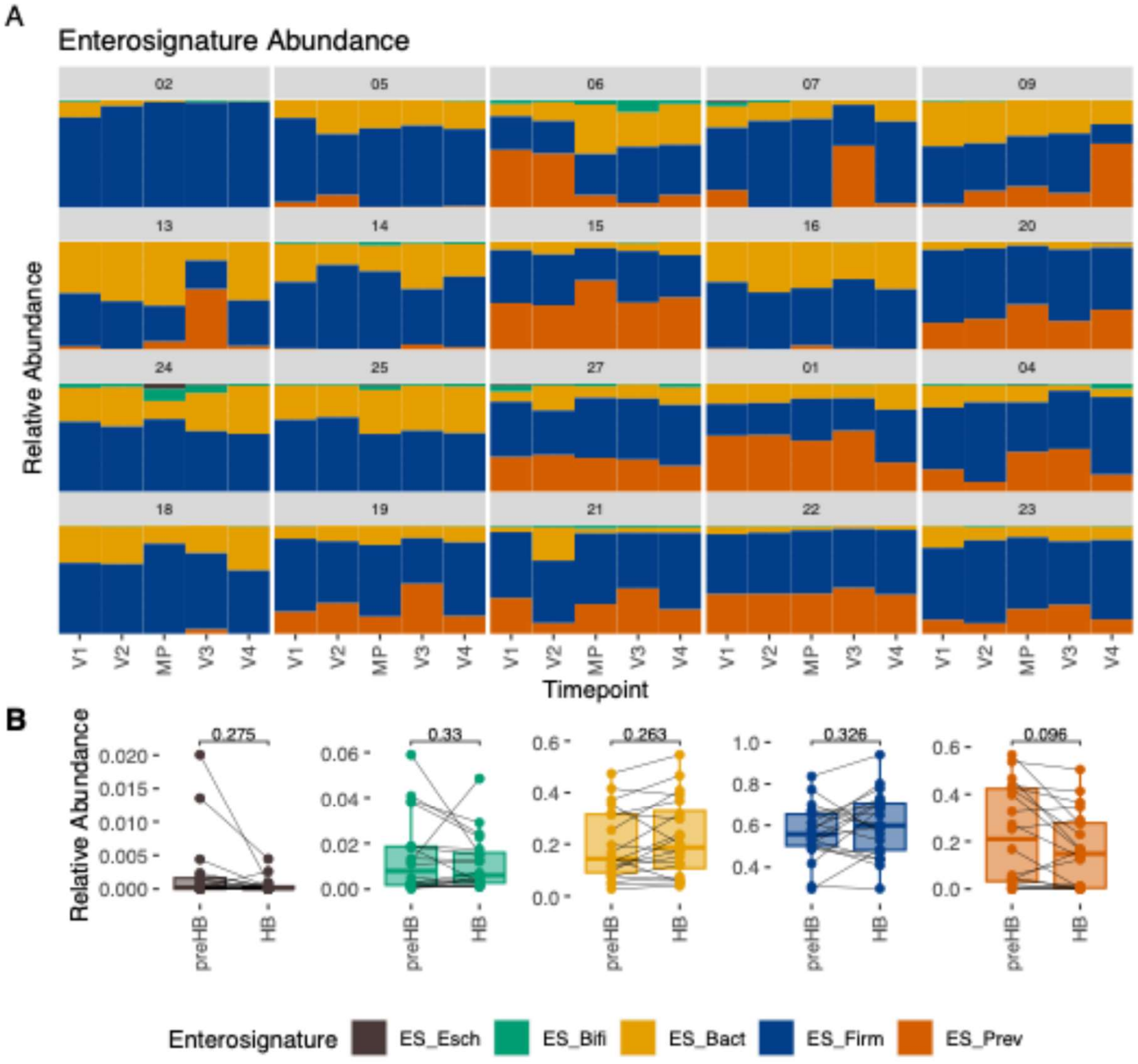


*Figure 7: A) Relative abundance of the five Enterosignatures (guild of bacterial genera) in each sample. Each block represents the samples for an individual participant, sorted by timepoint from first to last, left to right. B) Relative abundance of Enterosignatures comparing samples taken immediately before participants started the high bioactive diet (preHB) and immediately after the high bioactive diet (HB). Samples from the same participant are joined by lines. Significance tested with paired Wilcoxon test and Benjamini-Hochberg correction.*

#### dbRDA of Enterosignature weights with fitted fecal metabolites

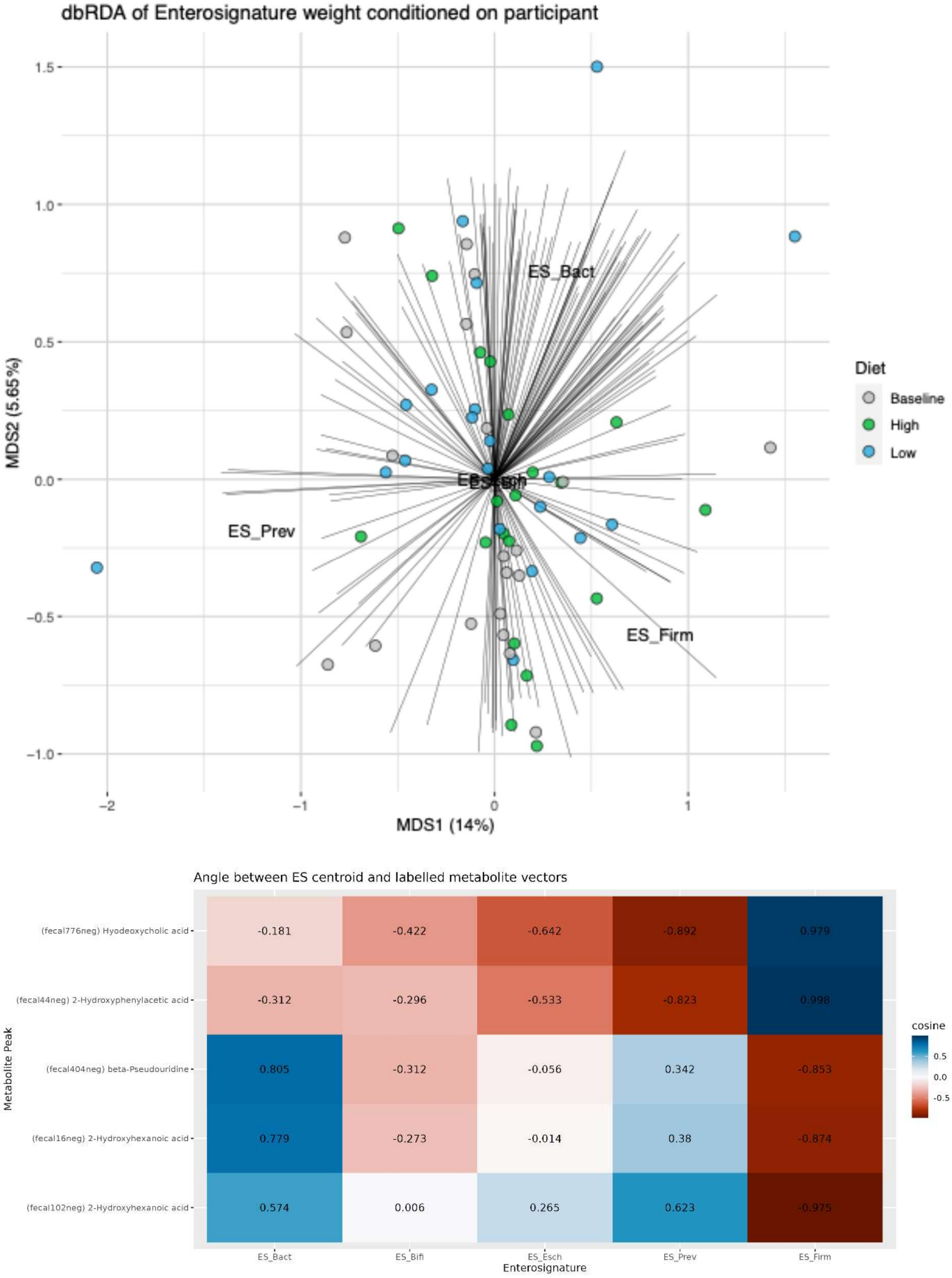


*Figure 8: Top: dbRDA of Enterosignature relative abundance, using Bray-Curtis dissimilarity. Participant as a conditioning term, which explained 80% of variance. Axes are labelled with the percentage of total variance explained (including that which is explained by the conditioning term). Fecal metabolite concentrations were fit to the ordination using the envfit function (perm=100), and only those with $p \leq 0.05$ shown, vectors are scaled by their correlation. Bottom: Angle between significant metabolite vectors which had identities assigned and Enterosignatures.*

#### Beta diversity between baseline and dietary interventions

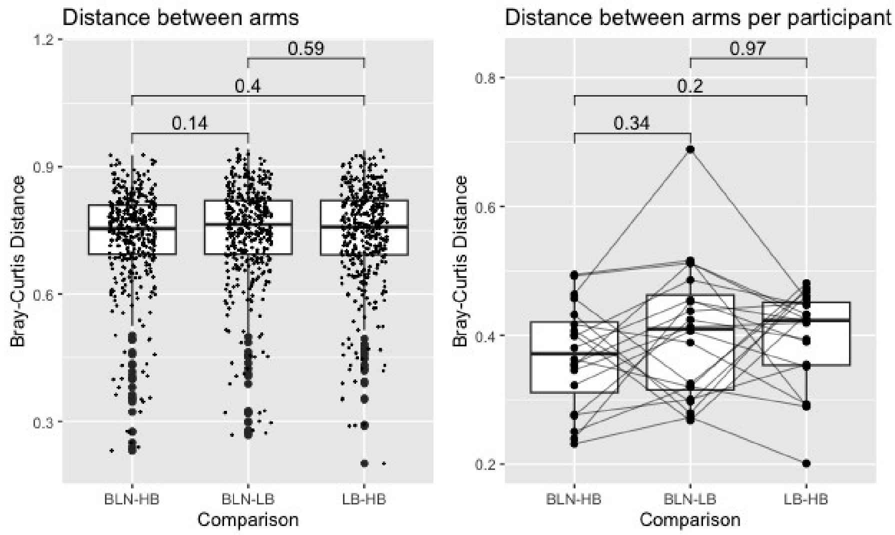


*Figure 9: Bray-Curtis Dissimilarity between samples from different study arms. Left shows all pairwise distances, difference tested with Wilcoxon rank-sum test. Right shows only distance between samples from the same participant, each participant joined by a line; difference tested with Wilcoxon rank-sum test.*

### Supplementary Results

#### Supplementary Results 1

The overall picture is that the features in HBcom-up and HBcom down in network analysis (Figure 4) are all related to medium-chain fatty acid metabolism but their role in fecal matter is elusive. Those in HBcom-up are related to dodecanedioic acid (free or as a carnitine) while those in HBcom-down most likely an O-tyrosyl esterified octenoylglycine; the octenoylglycine is known from mouse feces and urine, however its esterification to tyrosine has apparently never been described.

#### Identification for fecal1214pos m/z319.166_RT3.96

This is the second of two peaks with similar spectra, the former (and larger) is at 3.83min (Supplementary Figure 10). The feature is a daughter ion as documented by the parent ion (m/z 365.208) having a dimer at m/z 729.397 and a dimer with a sodium adduct (m/z 751.393). The sample with highest peak level is 22-2 (046 in sequence); the global pools have apparent higher intensities but that is an artifact based on inspection of the raw data. Daughter ions include m/z 319.166 (M-HCOOH), 291.172 (further loss of -C=O) , m/z 273.162 (further loss of water), and in the DDA-trace also m/z 166.086 (phenylalanine; loss of 199.122 (most likely 2-octenoylglycine)), m/z 148.074 (further loss of water), and 133.089 (further loss of NH); all of these are relatively a bit higher in the DDA runs, while the parent is very small. This fragmentation would indicate that the most likely structure of the compound is an ester of tyrosine and 2-octenoylglycine; 2-octenoylglycine has been reported in mouse feces (metabolights record), and may also be observed as a small and broad peak here (RT 3.64). It is also the most abundant short/medium-chain fatty acyl glycine in mouse urine (https://pubs.acs.org/doi/10.1021/acs.analchem.2c02507).

The compound is also observed in NEG mode, see further discussion below on compound NEG-703 and as shown in the chromatogram of positive and negative features and spectra below.

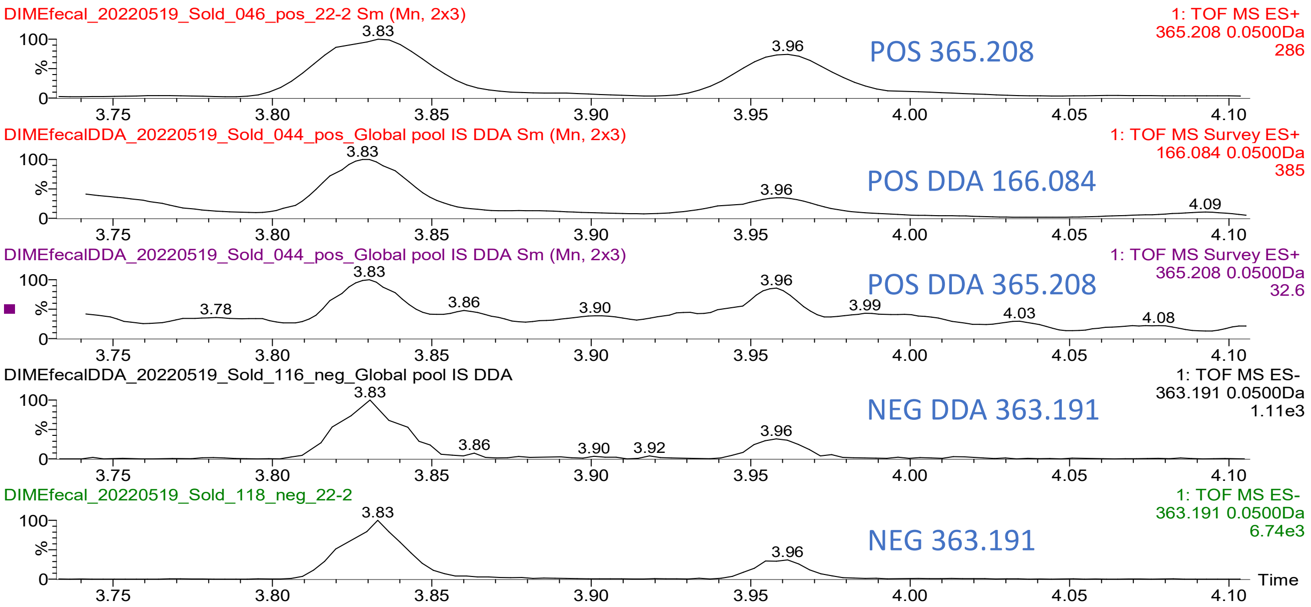

DIMEfecal_20220519_Sold_046_pos_22-2 Sm (Mn, 2x3)
1: TOF MS ES+ 365.208 0.0500Da 286
POS 365.208
DIMEfecalDDA_20220519_Sold_044_pos_Global pool IS DDA Sm (Mn, 2x3)
1: TOF MS Survey ES+ 166.084 0.0500Da 385
POS DDA 166.084
DIMEfecalDDA_20220519_Sold_044_pos_Global pool IS DDA Sm (Mn, 2x3)
1: TOF MS Survey ES+ 365.208 0.0500Da 32.6
POS DDA 365.208
DIMEfecalDDA_20220519_Sold_116_neg_Global pool IS DDA
1: TOF MS ES- 363.191 0.0500Da 1.11e3
NEG DDA 363.191
DIMEfecal_20220519_Sold_118_neg_22-2
1: TOF MS ES- 363.191 0.0500Da 6.74e3
NEG 363.191
Time


77

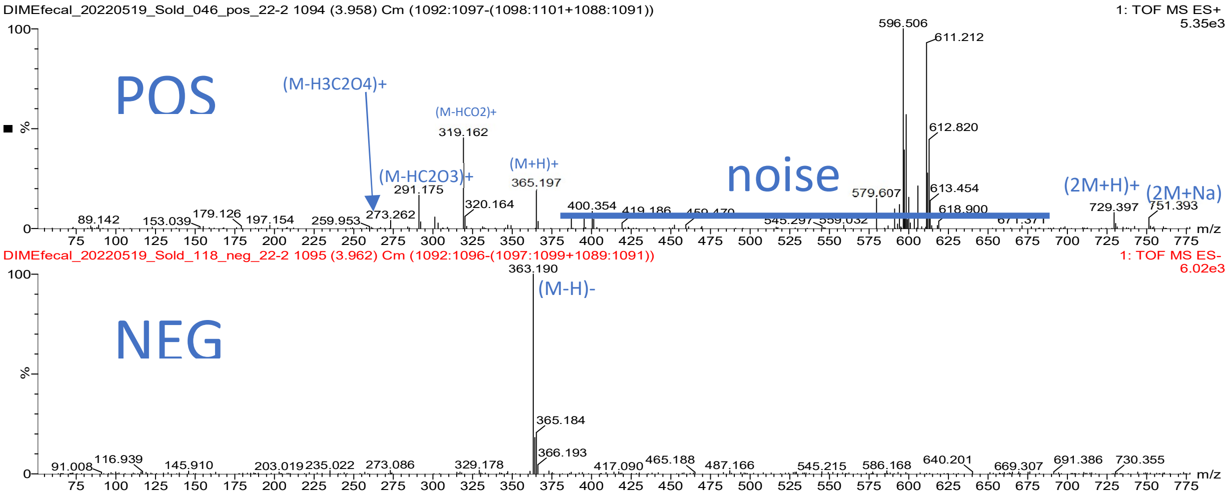

DIMEfecal_20220519_Sold_046_pos_22-2 1094 (3.958) Cm (1092:1097-(1098:1101+1088:1091))
1: TOF MS ES+ 5.35e3
POS
(M-H3C2O4)+
(M-HCO2)+
(M-HC2O3)+
(M+H)+
noise
(2M+H)+
(2M+Na)
DIMEfecal_20220519_Sold_118_neg_22-2 1095 (3.962) Cm (1092:1096-(1097:1099+1089:1091))
1: TOF MS ES- 6.02e3
NEG
(M-H)-


78

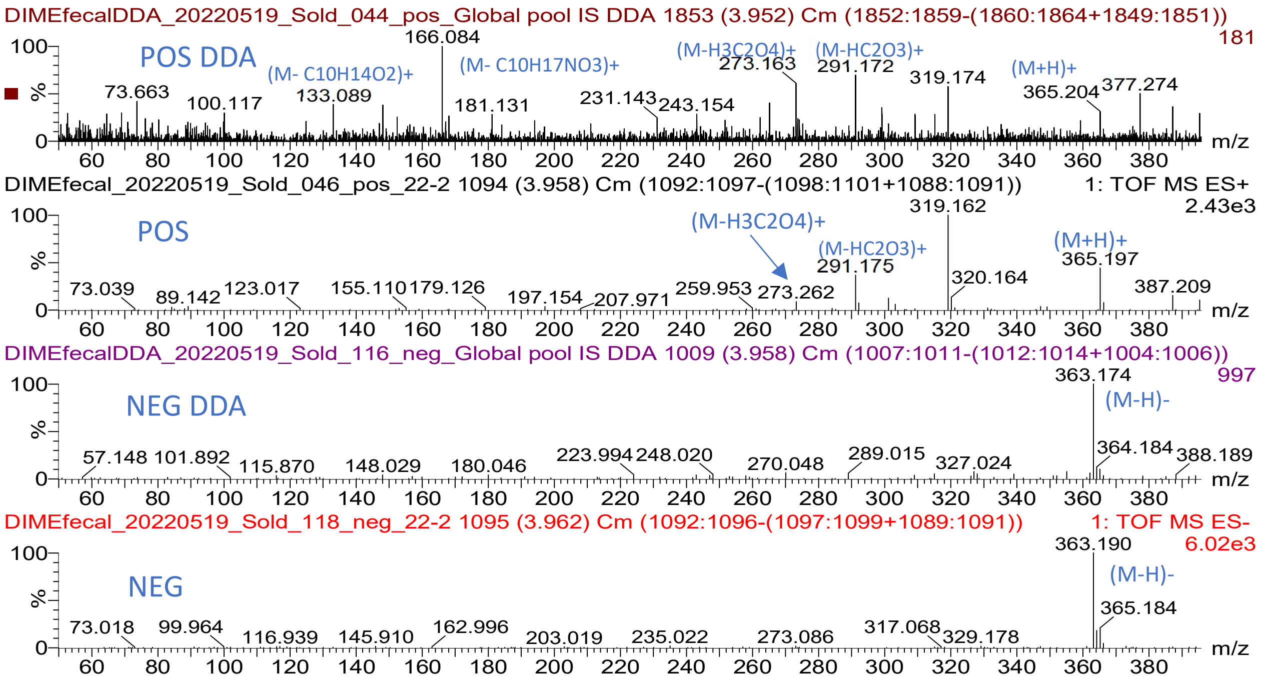

DIMEfecalDDA_20220519_Sold_044_pos_Global pool IS DDA 1853 (3.952) Cm (1852:1859-(1860:1864+1849:1851))
POS DDA
(M- C10H14O2)+
(M- C10H17NO3)+
(M-H3C2O4)+
(M-HC2O3)+
(M+H)+
DIMEfecal_20220519_Sold_046_pos_22-2 1094 (3.958) Cm (1092:1097-(1098:1101+1088:1091))
1: TOF MS ES+ 2.43e3
POS
DIMEfecalDDA_20220519_Sold_116_neg_Global pool IS DDA 1009 (3.958) Cm (1007:1011-(1012:1014+1004:1006))
NEG DDA
(M-H)-
DIMEfecal_20220519_Sold_118_neg_22-2 1095 (3.962) Cm (1092:1096-(1097:1099+1089:1091))
1: TOF MS ES- 6.02e3
NEG
(M-H)-
m/z


79

*Figure 10: Spectra for identification of fecal metabolite fecal1214pos.*

#### Identification for fecal703neg m/z363.1911_RT3.96

This peak like fecal POS-1214 is one of two with identical m/z and spectra, the larger eluting at 3.83 min. Very few, if any daughters or adducts can be observed in the negative mode spectrum in both the MS (samples 16-1 or 22-2, plate1) and DDA (global pool 116, plate 1) traces (Supplementary Figure 11) and no relevant compounds at this mass seem to be found in hmdb. In conclusion this feature is the same as POS-1214 but cannot be finally identified without further MS/MS analyses.

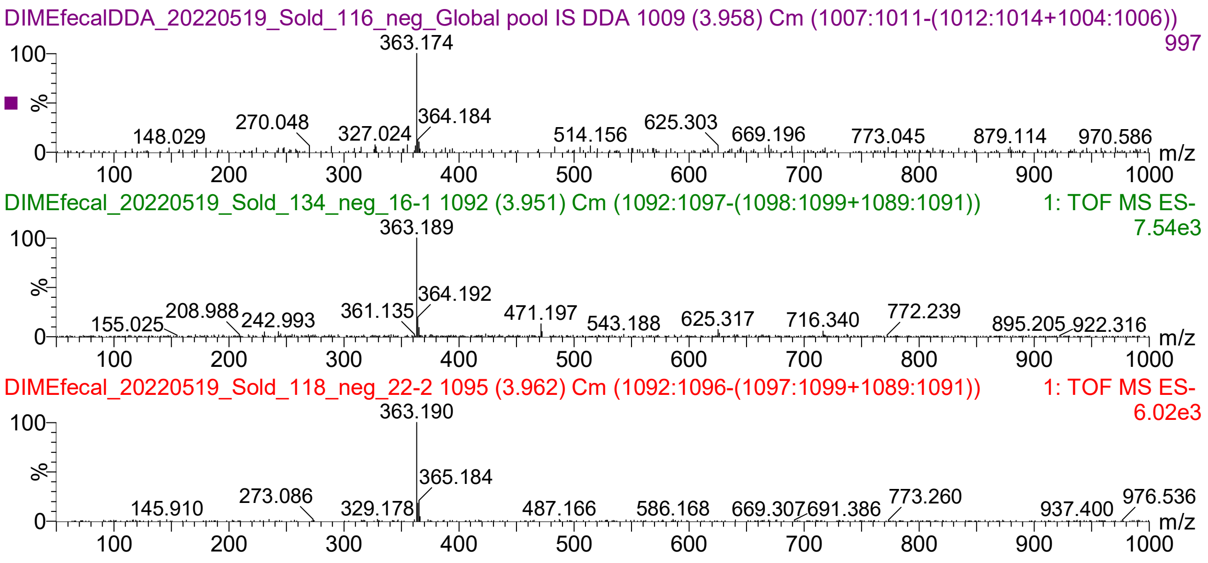


*Figure 11: Spectra for identification of fecal metabolite fecal703neg*

#### Identification for fecal535neg m/z299.1479_RT3.46

This is a very small peak. The largest peak is in sample 14-V2, but it shows no adducts or daughters. There is a peak with the same exact RT at 372.2142 (+73.0663, C4H9O) but this mass difference is not really useful, except that it might indicate that 372.2142 is the parent. The global pool is only 10% of that sample and in the DDA trace of the pool, the peak is absent. There are no plausible hits in hmdb, except that 372.2142 might be dodecanedioylcarnitine, but in that case it is 67ppm off (m/z 372.239). The corresponding peak (m/z 301.162) is extremely small in POS mode, and the characteristic daughter with m/z 85.1 is missing, but dodecanedioylcarnitine has the same RT in NEG mode (very small, uncertain peak) and a possible shows 85.063 daughter in NEG mode. The most likely identity is dodecanedioylcarnitine, however it should be synthesized to investigate whether it correlates with the target.

#### Identification for fecal367neg m/z227.127_RT4.02

This feature is very small in most samples, but again relatively high in sample 14-V2. It is a daughter (or has adducts) of features with mass 345.208 and 468.310 and may have additional daughters at 183.130 (loss of -COO) and 112.983 (further loss of -C=CH-COOH). The most likely composition of the 227-peak is C12H11O4 (M-H-, some 3ppm off), a dodecanedioic acid such as traumatic acid. The difference to 345.208 fits betaine or a dipropylsulfide  (both 20 ppm off) but neither 345.210 nor 468.314 matches anything plausible in relation to a loss of dodecanedioic acid. In DDA the 227-feature is reduced while all the others mentioned increase, except 183.129, which is gone. The 345- and 468-features shows very similar peak shapes with 227 in DDA, however as they are not affected by the energy, they must be independent features being there by chance. As seen in the figure below the retention time observed fits exactly with the predicted RT for dodecenedioic acid, based onother dicarboxylic aliphatic acids from C8-C14 observed in the same sample, however C12:0 and C12:1 are the most abundant. C12:0 has the same daughter at 112.983 but loses water before losing a carboxylic acid and then CH2-CH2-OH. The spectrum has only partial overlap with the predicted spectrum at hmdb.ca, both showing loss of HCOOH. Given the observed loss of two carboxylic acid functions, one of which is next to a double bond and the similarity with the spectrum of C12:0 (see annotated spectra below), the conclusion is that the compound is dodec-2-enedioic acid, which is identical to the plant hormone, traumatic acid.

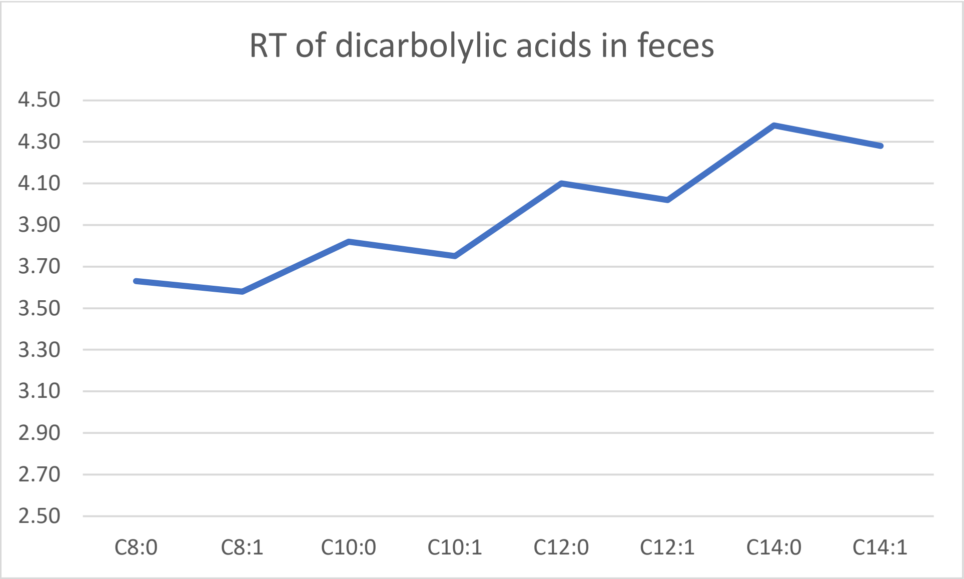

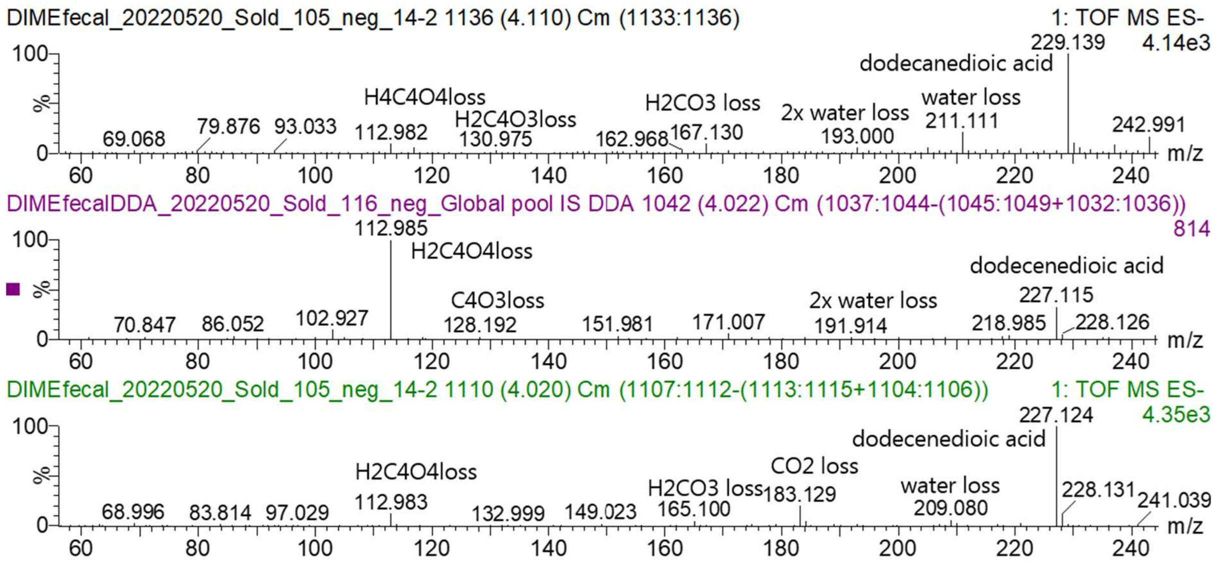


*Figure 12: Spectra for identification of fecal metabolite fecal367neg*

### Supplementary Tables

#### Demographic and clinical characteristics stratified by intervention

| **Clinical Measures** | **High Bioactives** | **Low Bioactives** |
|---|---|---|
| Weight (kg) | | |
| *Pre-intervention* | 73.1 ± 3.3 | 73.6 ± 3.3 |
| *Post-intervention* | 72.7 ± 3.2 | 72.9 ± 3.2 |
| Body Mass Index (kg/m$^2$) | | |
| *Pre-intervention* | 24.6 ± 0.9 | 24.8 ± 0.9 |
| *Post-intervention* | 24.5 ± 0.9 | 24.6 ± 0.9 |
| Waist Circumference (cm) | | |
| *Pre-intervention* | 85.6 ± 2.3 | 84.8 ± 2.4 |
| *Post-intervention* | 84.5 ± 2.4 | 85.5 ± 2.4 |
| Blood Pressure (mmHg) | | |
| *Pre-intervention* | 118/78 ± 2/2 | 118/79 ± 4/2 |
| *Post-intervention* | 118/75 ± 2/2 | 120/75 ± 4/2 |
| Fasting Blood Glucose (mmol/L) | | |
| *Pre-intervention* | 5.0 ± 0.1* | 5.3 ± 0.1 |
| *Post-intervention* | 5.4 ± 0.1* | 5.3 ± 0.1 |
| Total Cholesterol (mmol/L) | | |
| *Pre-intervention* | 5.0 ± 0.2 | 5.3 ± 0.2 |
| *Post-intervention* | 5.2 ± 0.2 | 5.2 ± 0.2 |
| Low Density Cholesterol (mmol/L) | | |
| *Pre-intervention* | 3.0 ± 0.2 | 3.0 ± 0.1 |
| *Post-intervention* | 3.0 ± 0.2 | 3.0 ± 0.2 |
| High Density Cholesterol (mmol/L) | | |
| *Pre-intervention* | 1.5 ± 0.1 | 1.5 ± 0.1 |
| *Post-intervention* | 1.5 ± 0.1 | 1.5 ± 0.1 |
| Triglycerides (mmol/L) | | |
| *Pre-intervention* | 1.3 ± 0.1 | 2.0 ± 0.5 |
| *Post-intervention* | 1.5 ± 0.2 | 1.4 ± 0.2 |

| | | |
|---|---|---|
| High Sensitivity C-Reactive Protein (mg/L) | | |
| *Pre-intervention* | 1.0 ± 0.2 | 1.3 ± 0.5 |
| *Post-intervention* | 1.5 ± 0.7 | 0.9 ± 0.2 |
| Mean ± SEM (standard error of mean)<br>*Pre and post intervention; student's t test (paired t-test, significant difference p≤ 0.01) | | |

*Table 1: Demographic and clinical characteristics of participants before and after each two week dietary intervention.*

#### Dietary intake during Baseline, High Bioactive, and Low Bioactive diet periods

| **Macronutrients** | **Baseline** | **High Bioactives** | **Low Bioactives** |
|---|---|---|---|
| Energy (kcal) | 1934 ± 49<br>(1551 to 4073) | 2156 ± 43<br>(1669 to 6818) | 2177 ± 35<br>(1765 to 4739) |
| Energy (kJ) | 8119 ± 205<br>(6449 to 17098) | 9150 ± 218<br>(7017 to 42634) | 9128 ± 149<br>(7390 to 19858) |
| Total Carbohydrates (g) | 217 ± 6.5<br>(166 to 557) | 245 ± 7.7<br>(180 to 1433) | 222 ± 4.0<br>(173 to 477) |
| Total Sugars (g) | 80 ± 3.1<br>(54 to 310) | 109 ± 3.4*<br>(74 to 495) | 82 ± 2.3*<br>(56 to 281) |
| *Oligosaccharide (g)* | 0.7 ± 0.9<br>(0 to 18.9) | 0.8 ± 0.1*<br>(0 to 12.1) | 0.3 ± 0.9*<br>(0 to 4.0) |
| *Sucrose (g)* | 16.8 ± 1.4<br>(6.3 to 135) | 22.2 ± 1.6*<br>(7.5 to 163) | 16.7 ± 0.1*<br>(6.5 to 105) |
| *Glucose (g)* | 10.4 ± 0.6<br>(4.8 to 49) | 17.1 ± 0.7*<br>(9.6 to 105) | 13.4 ± 0.7*<br>(5.3 to 83) |
| *Fructose (g)* | 12.3 ± 0.7<br>(5.0 to 49) | 19.9 ± 0.9*<br>(10.5 to 138) | 14.0 ± 0.7*<br>(5.1 to 80) |
| Dietary Fibre (g) | 23.4 ± 0.8<br>(17.3 to 68.2) | 31.4 ± 0.7*<br>(27.3 to 175) | 27.1 ± 0.4*<br>(24.1 to 50.6) |
| Total Protein (g) | 85 ± 2<br>(65 to 226) | 93 ± 2<br>(67 to 361) | 96 ± 2<br>(72 to 248) |
| Total Fat (g) | 75 ± 2<br>(56 to 194) | 80 ± 6*<br>(59 to 327) | 96 ± 2*<br>(69 to 231) |
| *Saturated Fat (g)* | 26 ± 1.0<br>(18 to 70) | 32 ± 1.1<br>(19 to 155) | 33 ±1.0<br>(21 to 81) |
| *Monounsaturated Fat (g)* | 21 ± 1.1<br>(12 to 93) | 23 ± 0.9*<br>(13 to 82) | 31 ± 1.0*<br>(18 to 93) |
| *Polyunsaturated Fat (g)* | 9.8 ± 0.6<br>(5.3 to 40) | 10.5 ± 0.4*<br>(5.5 to 42) | 12.7 ± 0.5*<br>(6.6 to 41) |
| **Micronutrients** | | | |
| Vitamin A (µg, total RE) | 997 ± 89<br>(299 to 6377) | 997 ± 86*<br>(352 to 19701) | 440 ± 86*<br>(168 to 17142) |
| Vitamin K1 (µg) | 62 ± 8.9<br>(7.5 to 2715) | 102 ± 9.1*<br>(12 to 1355) | 38 ± 2.9*<br>(6.1 to 335) |
| Total Folates (µg) | 249 ± 11.6<br>(150 to 851) | 288 ± 15.1*<br>(187 to 3509) | 222 ± 5.9*<br>(155 to 695) |
| Vitamin C | 99 ± 7.3<br>(36.9 to 581) | 183 ± 7.0*<br>(104 to 783) | 66.8 ± 3.5*<br>(30.7 to 523) |
| Magnesium (mg) | 292 ± 11.6<br>(202 to 804) | 294 ± 7.4*<br>(224 to 1018) | 335 ± 7.6*<br>(1243 to 928) |
| Copper (mg) | 1.10 ± 0.06<br>(0.63 to 4.11) | 1.15 ± 0.03*<br>(0.80 to 4.88) | 1.47 ± 0.07*<br>(0.93 to 12.5) |
| **Bioactive Compounds** | | | |
| Anthocyanins (mg) | 105 ± 19.1<br>(0 to 1168) | 260 ± 25.5*<br>(3151 to 17423) | 89 ± 14.4*<br>(0 to 140) |
| Beta carotene (µg) | 3542 ± 423<br>(266 to 42312) | 3545 ± 259*<br>(620 to 22541) | 358 ± 27.8*<br>(124 to 5294) |

| | | | |
|---|---|---|---|
| Caffeine (mg) | 122 ± 10.3<br>(50 to 806) | 87 ± 4.6*<br>(7 to 356) | 76 ± 4.1*<br>(0 to 403) |
| Ellagitannins/Ellagic Acid (mg) | 11.1 ± 2.6<br>(0 to 212) | 63.5 ± 3.9*<br>(16.5 to 381) | 0.1 ± 0.1*<br>(0 to 23) |
| Glucosinolates (mg) | 159 ± 28.7<br>(0 to 70) | 239 ± 30.9*<br>(0 to 3630) | 4.9 ± 2.9*<br>(0 to 704) |
| Lignans (mg) | 0.8 ± 0.1<br>(0.1 to 8.5) | 2.7 ± 0.2*<br>(0.6 to 18.0) | 0.7 ± 0.7*<br>(0.1 to 7.1) |

Mean ± SEM (standard error of mean)
RE: retinol equivalents
*High versus low bioactives intervention; student's t test (paired t-test, significant difference p≤ 0.01)

*Table 2: Intake of macro- and micronutrients during baseline period, and two week interventions. Intake was compared for all micronutrients and bioactives between high and low interventions, with only statistically significant results (p≤ 0.01) displayed in the table.*
